# EsoDetect: Computational Validation and Algorithm Development of a Novel Diagnostic and Prognostic Tool for Dysplasia in Barrett’s Esophagus

**DOI:** 10.1101/2024.11.26.24317976

**Authors:** Migla Miskinyte, Benilde Pondeca, José B. Pereira-Leal, Joana Cardoso

## Abstract

Barrett’s esophagus (BE) is the only known precursor to esophageal adenocarcinoma (EAC), a malignancy with increasing incidence and unfavorable prognosis. This study endeavors to identify BE biomarkers capable of diagnosing low-grade dysplasia (LGD) in BE, as well as biomarkers that can predict the progression from BE to EAC to be subsequently integrated into diagnostic and prognostic algorithms.

Datasets containing gene expression data from metaplastic and dysplastic BE, as well as EAC tissue samples, were collected from public databases and used to explore gene expression patterns that differentiate between non-dysplastic (ND) and LGD BE (for diagnostic purposes) and between non-progressed and progressed BE (for prognostic purposes). Specifically, for the diagnostic application, three RNAseq datasets were employed, while for the prognostic application, nine microarray datasets were identified, and 25 previously described genes were validated. A Thresholding Function was applied to each gene to determine the optimal gene expression threshold for group differentiation. All analyzed genes were ranked based on the F1-score metrics. Following the identification of genes with superior performance, different classifiers were trained. Subsequently, the best algorithms for diagnostic and prognostic applications were selected.

In evaluating the value of gene expression for diagnosis and prognosis, the analyzed datasets allowed for the ranking of biomarkers, resulting in eighteen diagnostic genes and fifteen prognostic genes that were used for further algorithm development. Ultimately, a linear support vector machine algorithm incorporating ten genes was identified for diagnostic application, while a radial basis function support vector machine algorithm, also utilizing ten genes, was selected for prognostic prediction. Notably, both classifiers achieved recall and specificity scores exceeding 0.90.

The identified algorithms, along with their associated biomarkers, hold significant potential to aid in the early management of malignant progression of BE. Their strengths lie in their development using multiple independent datasets and their ability to demonstrate recall and specificity levels superior to those reported in the existing literature. Ongoing experimental and clinical validation is essential to further substantiate their utility and effectiveness, and to ensure that these tools can be reliably integrated into clinical practice to improve patient outcomes.

## INTRODUCTION

Barrett’s esophagus (BE) is characterized by the replacement of the normal squamous epithelium lining the lower esophagus with specialized columnar cells (intestinal metaplasia) [1–4]. This transformation occurs because of chronic gastroesophageal reflux disease (GERD) [1, 2] and exposure to stomach acid [3]. Approximately 10% of patients with GERD are likely to progress to a diagnosis of BE over 5 years [5]. Individuals with BE have a significantly increased risk of developing esophageal adenocarcinoma (EAC). Typically, the progression of EAC starts with GERD, followed by abnormal columnar cells characteristic of BE, which, over time, can progress to dysplasia and eventually become EAC. Despite BE’s role as a precursor to EAC, the exact risk factors associated with BE are still not fully understood but include age (≥ 60-70 years), male gender [6], tobacco use [7, 8], obesity [9], and hiatal hernia [10].

The clinical relevance of BE relies on its role as the sole known precursor lesion for EAC [1, 11]. This specific type of esophageal cancer constitutes already around two-thirds of all cases of esophageal cancer in high-income countries [12], with 85,700 new EAC cases estimated worldwide in 2020. Over the next two decades, a staggering 65% increase (equivalent to approximately 55,600 additional cases annually) is predicted [13]. EAC is a major problem because of its association with poor survival rates, one of the lowest in oncology. Post-diagnosis, EAC presents a 23% 5-year survival and a median survival of only 15 months [14], highlighting the need for efficient methods for EAC management. This low survival is mainly due to late diagnosis, limited treatment options, poor prognosis, high rate of early metastasis, and difficulties in early detection [15].

Due to the low progression rate of BE to EAC (estimates 0.1-0.5, reviewed by [16]), most BE patients never progress to cancer. However, GERD is becoming increasingly prevalent, with a global estimate of 783 million prevalent cases in 2019 [17]. Factors like population growth, aging, lifestyle changes, and improved living standards contribute to the rising incidence of GERD [18]. As BE is a complication of GERD and a significant risk factor for EAC, the increasing prevalence of GERD cases represents a menace to future management of EAC. The increased prevalence of GERD leads to a higher incidence of BE cases and pressures for BE screening and diagnosis, resulting in a significant economic burden for patients, families, health services, and society.

Currently, BE serves as a critical warning sign and its surveillance is essential for effective risk stratification. BE screening and surveillance methods involve endoscopic sampling of biopsies from four quadrants according to the Seattle biopsy protocol [19, 20] followed by histological analysis to classify detectable BE lesions as non-dysplastic (NDBE), indefinite for dysplasia (IND), low-grade dysplasia (LGD), or high-grade dysplasia (HGD) [11]. Limitations to the success of current strategies include but are not limited to, difficulties with endoscopic identification of dysplasia, biopsy sampling error, low interobserver reproducibility in histologic assessment of dysplasia among pathologists, lack of reliable biomarkers, access to specialized care and patient compliance [21]. Variability in the endoscopic and histologic assessment are commonly known issues: BE endoscopic/pathological management is time-consuming and depends on the clinical experience of the physicians involved in the endoscopic examination and/or histological analysis – who are mostly available in BE reference centers. For example, one meta-analysis reported up to 25% and 24% of EACs were respectively missed during surveillance or when the analysis was restricted to NDBE patients [22]. Regarding histological analysis, the inter-observer agreement among pathologists has been reported as only 58% when it comes to distinguishing normal esophagus from BE and was even lower (less than 50%) when diagnosing LGD in BE patients [23, 24]. The lack of agreement can become particularly problematic when many cases of BE are classified as IND (60% of dysplastic cases in a study by Alshelleh et al. [25]) and when the interobserver agreement is even poorer than for LGD [26]. There is emerging evidence that the addition of biomarkers to risk stratification models could increase BE diagnostic accuracy compared to current surveillance methods [27]. These biomarkers range from the incorporation of more clinical variables [28, 29] to molecular features such as genomic instability [30–34], gene expression patterns [35, 36], epigenetics [37, 38], and proteomics [39]. In addition to biomarkers, the recent emergence of artificial intelligence (AI) tools opens the prospect of improving the effectiveness of BE diagnosis and surveillance. A recent meta-analysis revealed that deep learning algorithms applied to endoscopy images in the surveillance of BE-related neoplasia are highly accurate (pooled sensitivity and specificity of 90.3% and 84.4%, respectively) in detecting early HGD/EAC [40], despite the absence of data for LGD. However, most diagnostic and prognostic tools (biomarkers, AI), still lack substantial validation in large patient cohorts, refraining from their usage in clinical practice [41]. In addition, the new tools available do not reach yet maximum performance. For example, when predicting the neoplastic progression to HGD/EAC, both TP53 staining and Tissue Cypher test demonstrate high specificity (86% and 82%, respectively) but to the detriment of low sensitivity/recall (49% and 55%, respectively) [reviewed by [42]].

While it is not yet clear whether regular surveillance surely leads to earlier detection of dysplasia and consequently to a decrease in mortality from EAC [43] surveillance is still the only recommended strategy for BE and EAC management. There is room for new diagnostic and prognostic tools to support clinicians when diagnosing BE dysplasia and segmenting patients based on the risk of BE progression to EAC.

The current study explores the diagnostic and prognostic value of gene expression patterns from BE tissue samples from public datasets in the context of BE. Envisioning its clinical applicability, it aims to identify biomarkers that can accurately identify dysplasia within BE lesions (diagnostic application) and biomarkers that can predict the progression to EAC (prognostic application). It is also intended to understand the individual and combined predictive value of each selected biomarker in both contexts through their implementation using machine learning algorithms.

## 2. MATERIALS AND METHODS

### 2.1. Dataset Search

An exhaustive search for public datasets containing gene expression data related to BE, including normal esophageal epithelium, NDBE, BE with different degrees of dysplasia (LGD and HGD) and EAC was performed in the following databases: Pubmed (https://pubmed.ncbi.nlm.nih.gov/), Gene Expression Omnibus (GEO, https://www.ncbi.nlm.nih.gov/geo/), Sequence Read Archive (SRA, https://www.ncbi.nlm.nih.gov/sra), and European Genome-Phenome Archive (https://ega-archive.org/). For the diagnostic application, the aim was to distinguish between NDBE and LGD BE. For the prognostic application, non-progressed BE (nonP-BE) and progressed BE (P-BE) data was studied. P-BE was defined as a BE adjacent to EAC. A summary of the methodology used is represented in Figure 1 and described in detail below.

**Figure 1.**
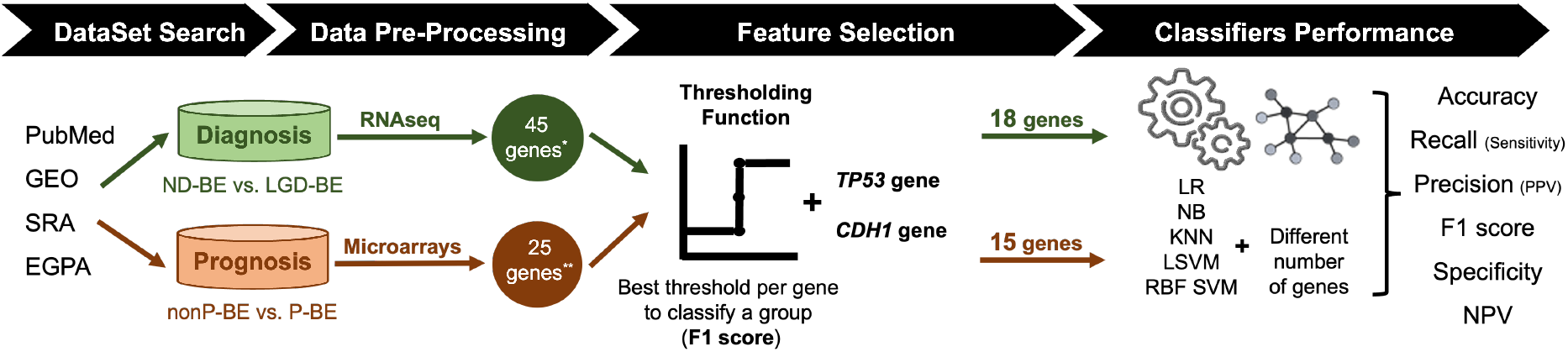
Methodology Summary. Datasets of interest were downloaded from public databases, such as PubMed, GEO (Gene Expression Omnibus), SRA (Sequence Read Archive), and EGPA (European Genome-Phenome). For the diagnostic application, i.e., the distinction between non-dysplastic (ND) BE and low-grade dysplasia (LGD) BE, RNAseq datasets were used. Low-expression genes were excluded, resulting in a pre-selection of 45 genes. For the prognostic application, i.e., the distinction between non-progressed Barrett’s Esophagus (nonP-BE) and progressed-BE (P-BE), microarray datasets were identified, and 25 previously described genes were selected [35]**. A Thresholding Function was applied to each gene to define the best gene expression threshold for group distinction. All analyzed genes were ranked by F1 score, and additional feature selection methods were applied for diagnostic genes*, determining the top genes for diagnosis and prognostic application. Due to their biological functions, two extra genes – *TP53* and *CDH1* – were added to both diagnostic and prognostic data sets, summing 18 diagnostic and 15 prognostic genes. Different algorithms – Logistic Regression (LR), Naive Bayes (NB), K-nearest neighbours (KNN), Linear Support Vector Machines (LSVM), and Radial Basis Function Support Vector Machines (RBF SVM) – were trained using different numbers of genes. Algorithms’ performance was assessed through accuracy, recall (sensitivity), precision (positive predictive value – PPV), F1 score, Specificity and negative predictive value (NPV). [35]

### 2.2. Data Pre-Processing

In this study, raw RNA-seq data from projects GSE193946, GSE58963, and E-MTAB-4054 were obtained from the Sequence Read Archive (SRA) and the European Genome-Phenome Archive (EGA). We processed the data using a Docker environment equipped with Kallisto version 0.46.1 (docker image: jlnetosci/kallisto:v0.46.1), which facilitated the pseudo-alignment of the reads against the Homo_sapiens.GRCh38.cdna.all.release-107 reference transcriptome from Ensembl. Post-alignment, the transcript abundance estimates generated by Kallisto were imported into the R programming environment using the tximport package. This allowed transcript-level data to be transformed into gene-level counts, which were subsequently analyzed for differential expression. The combined data was filtered for low-expressed genes using the filterByExpr function in EdgeR [44], resulting in a dataset of 20,608 genes for downstream analysis. Samples were then normalized using the TMM (Trimmed Mean of M-values) normalization method and differential expression analysis was performed using EdgeR [31]. For downstream analysis, including feature selection and classifier training, log-transformed CPM normalized values were used, which were subsequently corrected for batch effects using the ComBat function from the sva package [45].

In this study, microarray data was sourced from the Gene Expression Omnibus (GEO) database using the GEOquery package available in the R software. The data included accessions GSE1420, GSE363223, GSE13083, GSE37200, GSE34619, GSE26886, GSE39491, GSE100843, and an additional dataset from Watts et al. (2007) [46]. Data was loaded and normalized using both the affy and oligo packages in R, depending on the array platform. The CEL files were read and processed using the frma function for robust multi-array average (RMA) normalization.

Probe-level data was annotated and collapsed to gene-level data using Bioconductor annotation packages hugene10sttranscriptcluster.db, hgu133a.db, hgu133plus2.db, and hgu133a2.db along with the WGCNA package. Finally, the resulting gene expression data was merged into a single dataset for downstream analysis, with additional annotations indicating BE progression status [42][43]. For prognostic application, 25 genes selected in previous work to distinguish nonP-BE from P-BE [35], were used in this study – *ACTN1, C1S, CCN1* (alias *CYR61*), *CDH1, CEBPB, CEBPD, COL4A1, CTSB, DKK3, DUSP1, IER3, JUN, LAMC1, PLPP3, RBPMS, SNAI1, SNAI2, SPARC, TNS1, TRMT112, TP53, TWIST1, VWF, WWTR1* (alias *TAZ*) and *ZEB1*. Box plots representing normalized expression values were generated using the ggplot2 (v3.4.0) and ggsignif (v0.6.4) R packages. Statistical analysis was performed using one-way ANOVA, followed by a *post hoc* Tukey’s ‘Honest Significant Difference’ test, both from the R stats package (v4.1.1). When ANOVA assumptions were not met, a Kruskal-Wallis Rank Sum Test (R stats package v4.1.1) was performed, followed by a *post hoc* Dunn’s Kruskal-Wallis Multiple Comparisons test (FSA R package v0.9.3). The significance threshold was set at *p-value* < 0.05.

### 2.3. Threshold selection and determination of individual predictive power

For the distinction between NDBE and LGD (diagnostic) or nonP-BE and P-BE (prognostic) a Thresholding function was applied to the expression levels of each selected gene to determine an expression threshold. Performance metrics such as accuracy, recall (or sensitivity), precision (or positive predictive value – PPV), specificity, negative predictive value (NPV), and false positive rate (FPR) were calculated for each threshold, considering the known class of the samples. For the diagnostic application, other feature selection methods (Lasso, Mutual Information (MI) criteria, Recursive Feature Elimination (RFE), SelectKBest) were also applied to narrow down the most informative features that appeared at least twice in one of the methods. The threshold that yielded the highest F1-score was selected. Based on this metric, genes were ranked and the top 16 (diagnostic) and top 13 (prognostic) were considered for downstream analysis. Two additional genes – *TP53* and *CDH1* – were also included in the downstream analysis of both prognostic and diagnostic gene sets.

### 2.4. Algorithmic analysis and evaluation of performance metrics

Gene expression values were used for algorithm training. Several classes of classifiers, with shown applicability to microarray and RNAseq data [47–49], such as Logistic Regression (LR), Naive Bayes (NB), K-nearest neighbours (KNN), and Support Vector Machines (SVM) (with Linear and Radial Basis Function kernels), were implemented with default hyperparameters in Python programming language (v3.10.0), using the scikit-learn package (v1.0.1). A leave-one-out cross-validation procedure was used to evaluate the diagnostic or prognostic value of all possible combinations of genes (from n=2 up to all selected diagnostic or prognostic genes). This involved leaving out one sample at a time for validation while using the remaining samples to create a balanced training set. The Synthetic Minority Oversampling Technique (SMOTE) was employed from the imbalanced-learn (v0.8.1) package. For LR, KNN, and SVM, features were standardized (scaled and centered) using scikit-learn’s standard scaler module by subtracting the mean and scaling to the unit variance. Performance metrics such as accuracy, precision (PPV), recall (sensitivity), NPV, and precision and specificity were calculated and recorded for each full iteration of the validation strategy. The top-performing algorithms were chosen by maximizing performance metrics (accuracy, specificity, precision, recall, NPV, and F1-score, Table 2). The most frequent models, with the highest F1-score, were chosen to further select the best classifiers for both diagnostic and prognostic applications. The most frequently occurring genes (frequency ≥ 50 %) within the selected classifiers were chosen as features. Subsequently, the performance metrics were calculated using a decremental number of features, and the median value and standard deviation of each group of decremental subsets of genes were computed.

### 2.5. *In-vivo* gene expression analysis

#### 2.5.1. RNA extraction

Cell pellets from the cell lines metaplasia (BAR-T and BAR-T10 - from R. Souza, Baylor University Medical Center, Dallas, TX; Jaiswal et al., 2007; X. Zhang et al., 2010), dysplasia (CP-B, CP-C and CP-D - from P. Rabinovitch, University of Washington, Seattle, WA; Palanca-Wessels et al., 2003), and EAC (OE33, KYAE-1- from W. Dinjens, Erasmus Medical Center Cancer Institute, Rotterdam, Netherlands, and ESO26 - Boonstra et al., 2010) were used to extract RNA using the RNeasy Mini Kit (#74104, Qiagen, Hilden, Germany), following the manufacturer’s instructions.

For formalin-fixed paraffin-embedded (FFPE) tissue samples, RNA was isolated from 2 consecutive sections per sample, each approximately 20 mm2 and 5 μm. Tissue samples were deparaffinized using the deparaffinization solution (#19093, Qiagen, Hilden, Germany) prior to RNA extraction with the RNeasy FFPE Kit (#73504, Qiagen, Hilden, Germany), according to the manufacturer’s instructions (with one modification: proteinase K incubation was performed overnight).

All procedures involving human tissue samples were approved by the National Ethics Committee for Clinical Research – Comissão de Ética para a Investigação Clínica (CEIC), under approval number 2022_EO_24.

#### 2.5.2. Reverse Transcription - quantitative real-time Polymerase Chain Reaction (RT-qPCR)

For 1-Step RT-qPCR, reactions were performed in triplicate, using the TaqPath 1-step RT-qPCR Master Mix (#A15300, Thermo Fisher Scientific) with a final reaction volume of 10 μL. Each reaction containing 1 μL of template, 0.25 μM of probe and 0.5 μM of each primer. Data acquisition and analysis were conducted using the QuantStudio Design & Analysis Software v1.5.1 software, using the cycling program: UNG incubation at 25°C - 2 minutes, Reverse Transcription at 50°C - 15 minutes, followed by Polymerase activation at 95°C - 2 minutes and 40 cycles of Amplification at 95°C - 3 s and 58°C - 30 s. To normalize gene expression levels, the geometric mean of the reference genes (*PGK1*, *ELF1*, and *RPL13A*) was subtracted from cycle threshold (Cq) of the target genes.

## 3. RESULTS

### 3.1. Diagnosis and prognosis dataset selection

For the development of the diagnostic application, 13 RNAseq-based datasets were identified, of which only 3 had publicly available clinical data – GSE58963 [50], E-MTAB-4054 [51], GSE193946 [52] – and were therefore included in the present study. The BE data contained in each dataset is represented in Table 1. In total, data from 61 samples – comprising 21 NDBE, 40 LGD BE and 27 HGD – were included in the study.

**Table 1.**
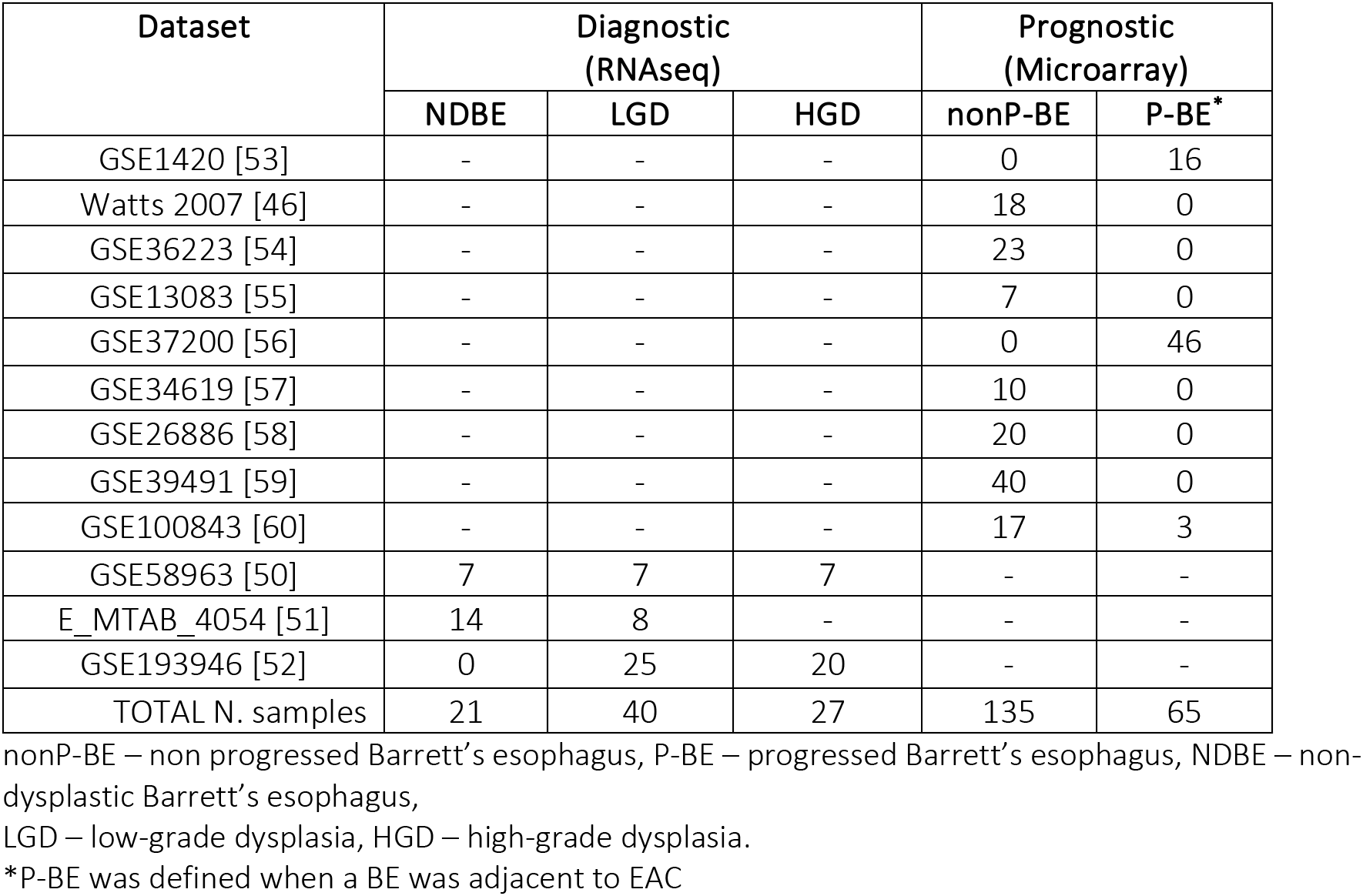
Characterization of datasets for the diagnostic and prognostic applications.

| Dataset | Diagnostic (RNAseq) |  |  | Prognostic (Microarray) |  |
| --- | --- | --- | --- | --- | --- |
|  | NDBE | LGD | HGD | nonP-BE | P-BE* |
| GSE1420 [53] | - | - | - | 0 | 16 |
| Watts 2007 [46] | - | - | - | 18 | 0 |
| GSE36223 [54] | - | - | - | 23 | 0 |
| GSE13083 [55] | - | - | - | 7 | 0 |
| GSE37200 [56] | - | - | - | 0 | 46 |
| GSE34619 [57] | - | - | - | 10 | 0 |
| GSE26886 [58] | - | - | - | 20 | 0 |
| GSE39491 [59] | - | - | - | 40 | 0 |
| GSE100843 [60] | - | - | - | 17 | 3 |
| GSE58963 [50] | 7 | 7 | 7 | - | - |
| E_MTAB_4054 [51] | 14 | 8 | - | - | - |
| GSE193946 [52] | 0 | 25 | 20 | - | - |
| TOTAL N. samples | 21 | 40 | 27 | 135 | 65 |

nonP-BE – non progressed Barrett’s esophagus, P-BE – progressed Barrett’s esophagus, NDBE – non-dysplastic Barrett’s esophagus,
LGD – low-grade dysplasia, HGD – high-grade dysplasia.
\*P-BE was defined when a BE was adjacent to EAC

For the prognostic application, 16 microarray datasets were identified, but only those generated on an Affymetrix platform were included in the downstream analysis to facilitate data merging. A total of 9 microarray datasets were analyzed, including three previously analyzed by Cardoso, *et al* [35], – GSE1420 [53], Watts_2007 [46], and GSE13083 [55] and six new ones, namely GSE36223 [54], GSE37200 [56], GSE34619 [57], GSE26886 [58], GSE39491 [59], and GSE100843 [60]. In total, data from 200 samples – representing 135 nonP-BE and 65 P-BE – were included in the study as shown in Table 1.

### 3.2. Identification of differentially expressed genes in a diagnostic and prognostic setting

In this study, we aimed to identify diagnostic biomarkers that can distinguish between ND-BE and LGD-BE. For this purpose, we utilized three RNAseq datasets (as listed in Table 1). Low-expression genes were excluded from each dataset, resulting in the inclusion of 20608 genes in our analysis. After normalization, we conducted differential expression analysis between LGDBE and NDBE (Figure S1A), and HGDBE and NDBE (Figure S1B) using EdgeR’s quasi-likelihood approach. This approach accounted for disease staging and batch effects from the three different datasets as factors in the model (Supplementary Table 1, Figure S1C).

Following the differential expression analysis, we identified 30 biomarkers through a systematic selection process. First, we selected differentially expressed genes (DEGs) with an absolute log fold change (logFC) of ≥ 1 between LGDBE and NDBE, with a false discovery rate (FDR) of < 0.05. From these DEGs, we filtered for genes that showed the same direction of expression change in the HGDBE vs. NDBE comparison (FDR < 0.05), resulting in 14 genes (Figure S1A). Second, we identified DEGs in the HGDBE vs. NDBE comparison with an absolute logFC of ≥ 2 (FDR < 0.05). Among these genes, we selected those that also exhibited the same direction of expression change in the LGDBE vs. NDBE comparison (considering p-value < 0.05 for significance), resulting in 16 genes (Figure S1B). This two-step filtering strategy ensured that the selected biomarkers not only had significant differential expression but also consistent expression patterns across different stages of disease progression. Given the strong batch effect observed (see Figure S1), there was a risk of losing biologically relevant genes in the LGDBE vs. NDBE comparison due to this variation. To mitigate this problem, we also performed separate analyses of the EMTAB_4054 (Supplementary Table 2) and GSE58963 (Supplementary Table 3) datasets. We employed the glmRobust pipeline to independently identify differentially expressed genes between the LGDBE and NDBE groups within each dataset. From these separate analyses, we identified an additional 13 genes with an absolute logFC greater than 1 and an FDR < 0.05. These genes were consistently found in both datasets and exhibited the same direction of expression change (Figure S2). Moreover, these genes showed consistent directional changes in the previous HGDBE vs. NDBE comparison. Thus, they were also included in the biomarker list (Supplementary Table 4). Given their established role in the biology of BE and EAC, we also included two additional genes – *TP53* and *CDH1* –in the downstream analysis, resulting in a total of 45 candidate genes for distinguishing between NDBE and LGD.

For the prognostic set of biomarkers, we re-analyzed 25 genes that we had previously identified to have prognostic value [35], namely *ACTN1, C1S, CCN1* (alias *CYR61*), *CDH1, CEBPB, CEBPD, COL4A1, CTSB, DKK3, DUSP1, IER3, JUN, LAMC1, PLPP3, RBPMS, SNAI1, SNAI2, SPARC, TNS1, TP53, TRMT112, TWIST1, VWF, WWTR1* (alias *TAZ*) and *ZEB1*. For validation purposes, we added six independent datasets to the three datasets we originally analyzed. We observed significant differential gene expression (adj. *p-value* < 0.05) between P-BE and nonP-BE categories for most of the genes of interest, except for *CDH1*, *DKK3*, *SNAI2, and WWTR1*.

### 3.3. Application of a Thresholding function for the selecting genes with the highest predictive value

To each selected gene, we applied a Thresholding function, to determine a gene expression threshold for distinguishing different levels of gene expression between groups of samples with distinct diagnosis (NDBE vs. LGD-BE) or with distinct prognosis (nonP-BE vs. P-BE). We defined the best individual threshold of gene expression for each selected gene, 45 for diagnosis and 25 for prognosis, reflecting the individual predictive value of each gene. Genes were then ranked by the harmonic mean of recall and precision (F1-score) to ensure accurate selection. This procedure identified the top 15 genes for predicting the malignant progression of BE lesions with F1-score above 0.67 (Supplementary Table 5). From the top 45 diagnostic genes, including CDH1 and TP53, genes with higher expression values (log2CPM above 1) were filtered. To further refine a list of candidates, we used several feature selection methods: Lasso, Mutual Information (MI) criteria, Recursive Feature Elimination (RFE), and SelectKBest. Additionally, feature correlation analysis was conducted to identify and eliminate highly correlated features (Pearson’s correlation coefficient > 0.9). Hence, for diagnostic purposes, we further narrowed down the selection to genes that were chosen at least twice in one of the feature selection methods and F1-score above 0.7, which identified the top 16 genes for diagnosing dysplasia in the context of BE (Supplementary Table 6).

Building on our identification of the top diagnostic and top prognostic genes using the Thresholding function and various feature selection methods, the ROC curve results (Figure 2) further validate their predictive power using a logistic regression classifier. In both diagnostic (Figure 2A) and prognostic (Figure 2B) contexts, most of the genes have AUC (Area Under the Curve) values above 0.50 (random chance line), with some reaching individual values of 0.90, demonstrating a substantial predictive value of the selected genes in both diagnostic and prognostic applications.

**Figure 2.**
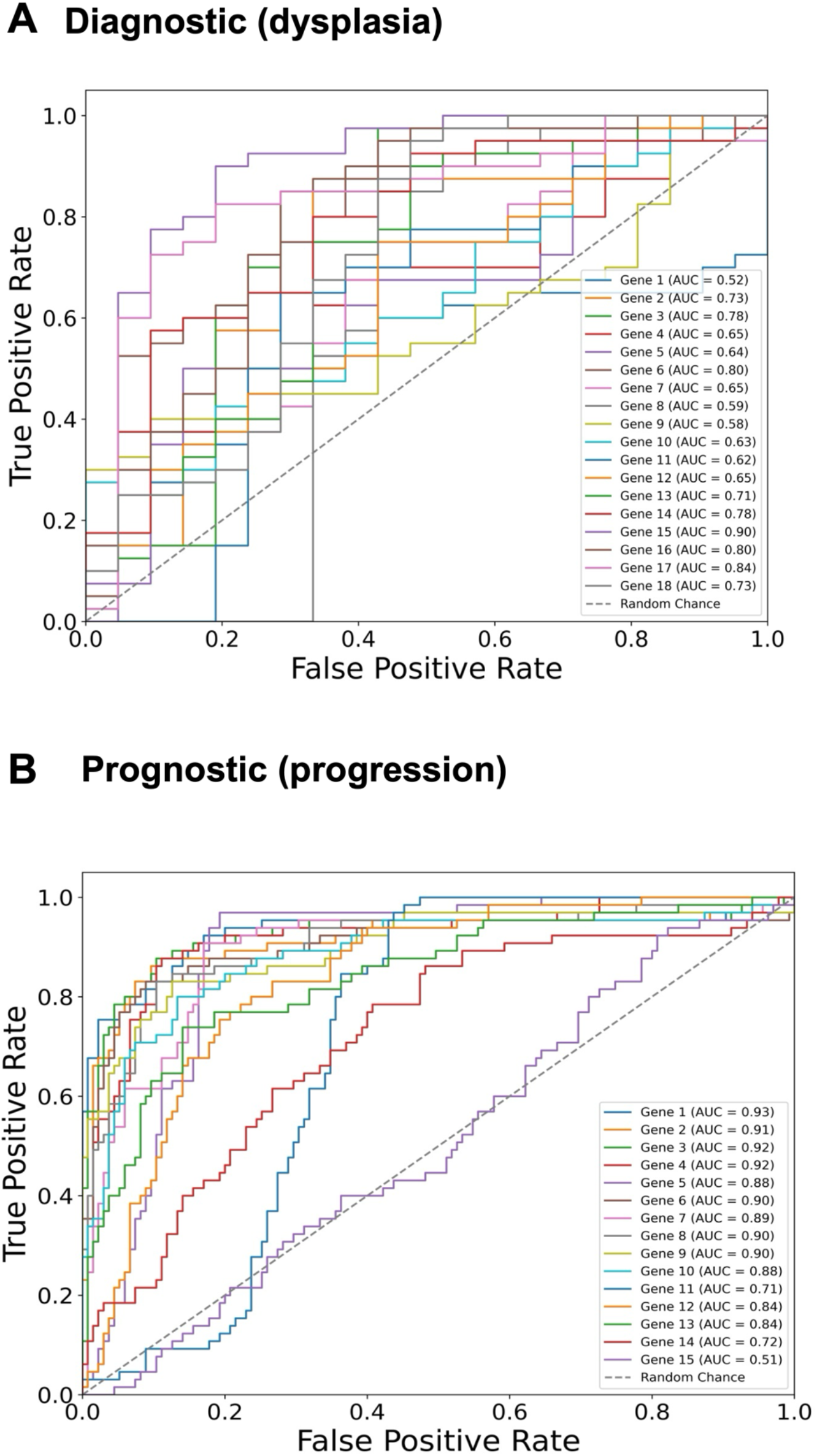
Gene-Specific ROC Curves for Diagnostic and Prognostic Predictions. Receiver Operating Characteristic (ROC) curves for individual genes depicting their predictive value in two contexts: (A) Diagnostic (dysplasia) and (B) Prognostic (progression) using a logistic regression classifier. The Area Under the Curve (AUC) values for each gene are indicated in the legends. Notably, the predictive values of TP53 and CDH1 genes are also included, although they were manually added to the sets.

### 3.4. SVM algorithms were the best for both diagnostic and prognostic applications

The diagnostic and prognostic gene groups were utilized to train the most effective diagnostic and prognostic algorithms. Various classifiers – LR, NB, KNN, LSVM, and RBF SVM – were examined using increasing combinations of genes, ranging from n = 2 up to the total number, for diagnostic and prognostic applications.

The algorithms were ranked based on their performance metrics for each application (see Table 2). However, no algorithms optimized all performance metrics for both applications. Nevertheless, the LSVM algorithms emerged as the best for diagnostic purposes, maximizing the F1-score and accuracy (refer to Figure 3A and Table 2).

**Table 2.**
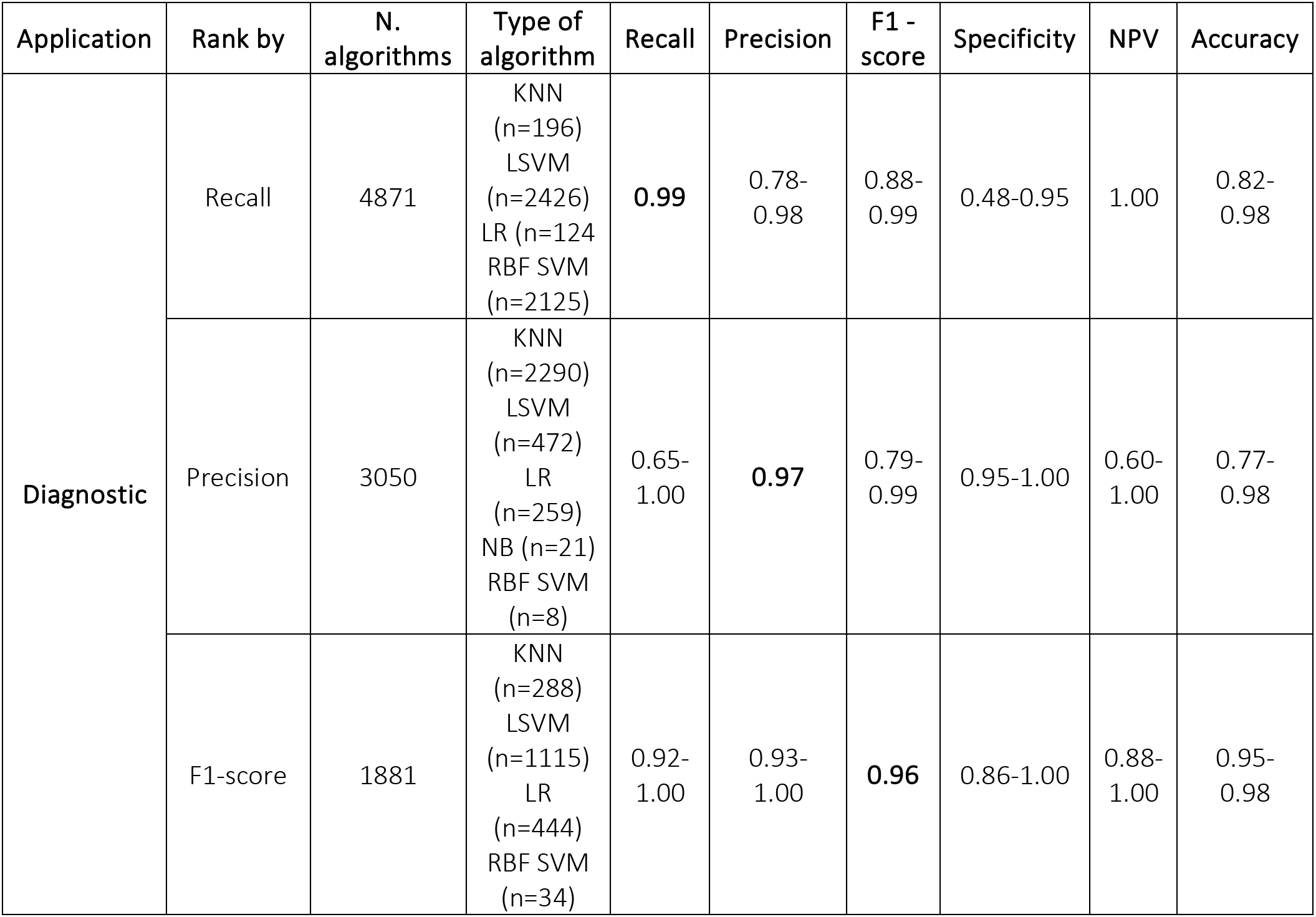
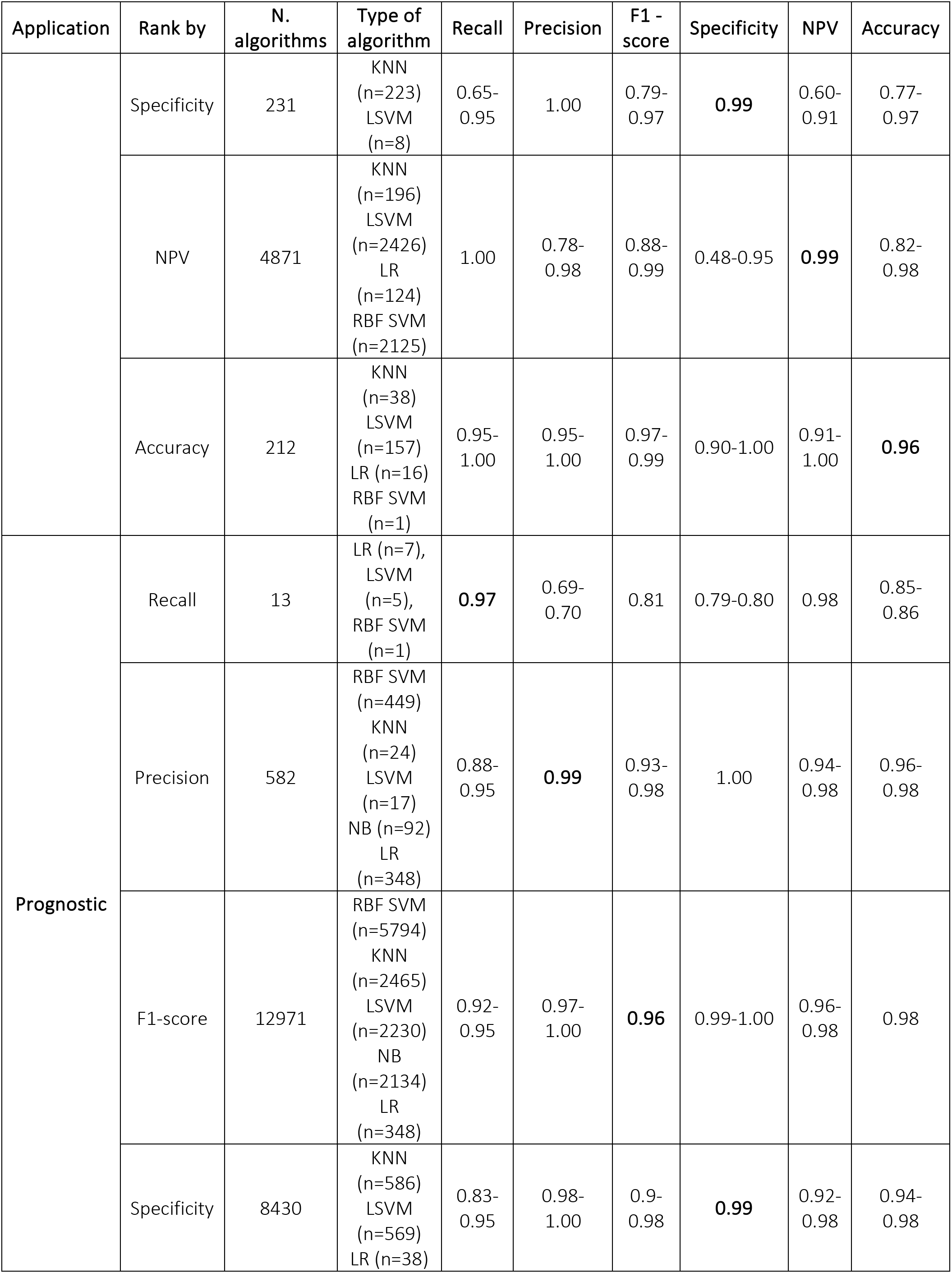
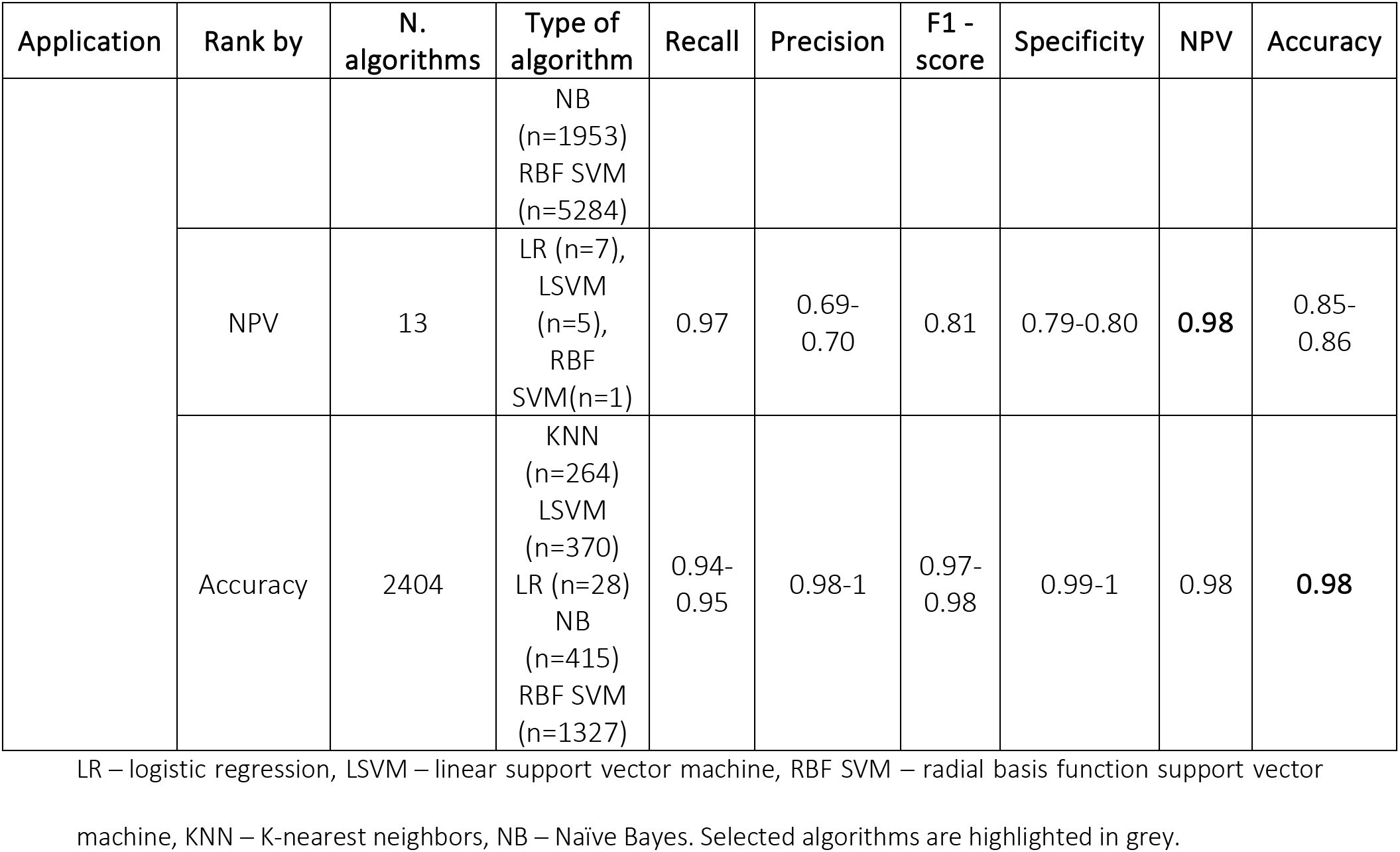
Best algorithm performance by metric maximization.

**Figure 3.**
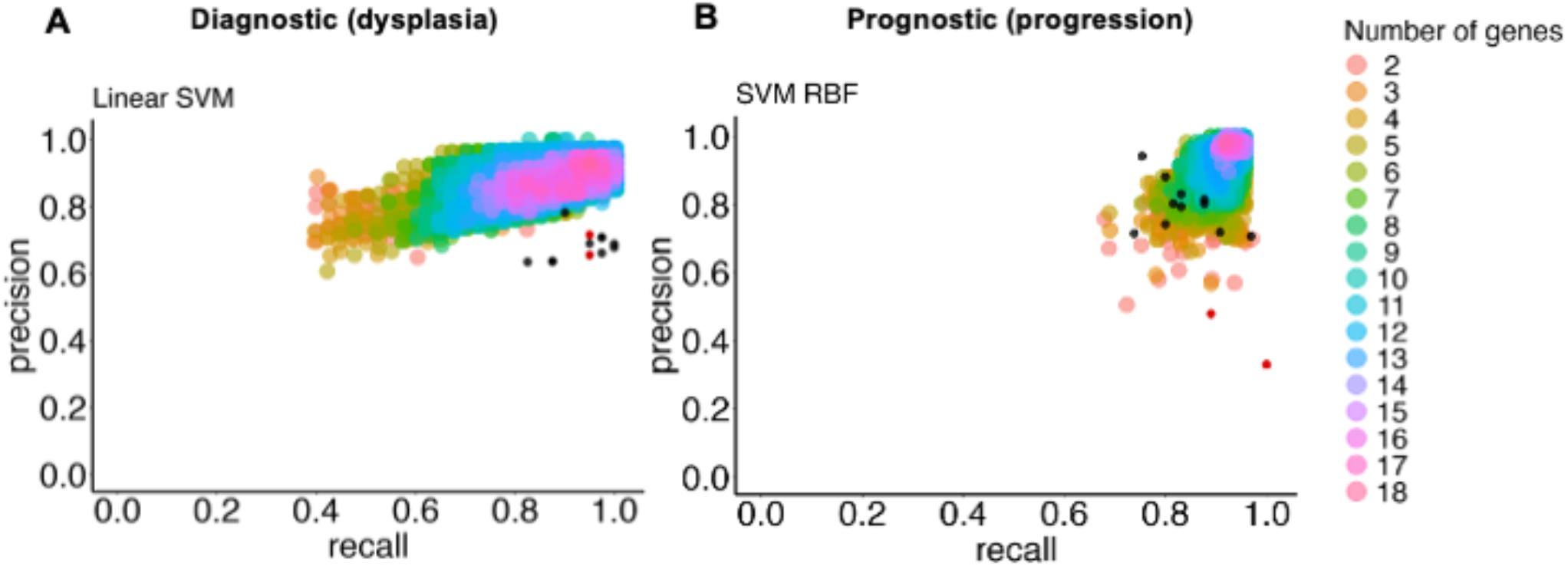
Precision and Recall for the selected classifier type with increasing combinations of genes of interest to predict BE dysplasia (diagnostic) and BE malignant progression (prognostic). This illustrates the performance of the chosen classifier types in the predicting dysplasia (A) and progression (B) when different numbers of genes of interest are combined (colored dots). The individual predictive value for the best threshold of each previously selected gene is also represented (black dots) for diagnostic (see Figure 3A) and prognostic (see Figure 3B). Colors represent different numbers of combined genes. LSVM – Linear Support Vector Machine (A), and RBF SVM – Radial Basis Function Support Vector Machine (B). Red dots represent manually added CDH1 and TP53 genes.

For the prognostic application, a similar trend was observed, where the RBF SVM type performed best according to the F1-score and accuracy metrics (refer to Figure 3B and Table 2).

The study found that among the selected types of algorithms, those with an F1-score above 0.96 included 1115 LSVM for diagnostic and 5794 RBF SVM for prognostic. The analysis identified the most frequent genes (over 50 %) across the best-performing algorithm class (Supplementary Table 7 and Supplementary Table 8). Ultimately, ten genes were selected for identifying LGD BE using a LSVM algorithm: *IGHV3-43, SLC38A4, PLLP, CELA3A, IGHV4-31, TMPRSS5, TP53, NR4A1, ATF3, IFI27*. For identifying P-BE, ten genes were selected using an RBF SVM algorithm: *SNAI1, C1S, DUSP1, CEBPB, COL4A1, ZEB1, CEBPD, CCN1, LAMC1* and *TWIST1*.

The performance of each selected algorithm (LSVM for diagnosis and RBF SVM for prognosis) was evaluated using the most frequent genes (10 for diagnosis and 10 for prognosis) as features. To test different random states while avoiding algorithm bias, 100 runs were performed for each algorithm with the same features. Table 3 presents the mean values and respective standard deviations (SD) for each performance metric. All performance metrics were above 0.90, except for specificity for the LSVM diagnostic algorithm. The low standard deviations (below 0.05) indicated an increase in the predictive value of each algorithm when the selected genes were combined.

**Table 3.** Performance of the selected algorithms with the selected genes as features after 100 runs.

A. Diagnostic
|  | Mean | Standard deviation |
| --- | --- | --- |
| Accuracy | 0,946 | 0,014 |
| Precision | 0,932 | 0,012 |
| Recall | 0,991 | 0,017 |
| F1 score | 0,960 | 0,010 |
| TP | 39,630 | 0,677 |
| FP | 2,900 | 0,541 |
| TN | 18,100 | 0,541 |
| FN | 0,370 | 0,677 |
| NPV | 0,981 | 0,033 |
| Specificity | 0,862 | 0,026 |
| FPR | 0,138 | 0,026 |

B. Prognostic
|  | Mean | Standard deviation |
| --- | --- | --- |
| Accuracy | 0,977 | 0,003 |
| Precision | 0,977 | 0,008 |
| Recall | 0,952 | 0,005 |
| F1 score | 0,965 | 0,004 |
| TP | 61,900 | 0,302 |
| FP | 1,430 | 0,498 |
| TN | 133,570 | 0,498 |
| FN | 3,100 | 0,302 |
| NPV | 0,977 | 0,002 |
| Specificity | 0,989 | 0,004 |
| FPR | 0,011 | 0,004 |

TP- True Positive, FP- False Positive, TN- True Negative, FN- False Negative, NPV- Negative Predictive Value, FPR- False Positive Rate.

Finally, the performance of the two algorithms was evaluated by gradually decreasing the number of selected genes (Figure 4). The diagnostic algorithm showed a decrease in performance after the removal of just one gene (Figure 4A). In contrast, the prognostic algorithm showed noticeable changes only after the removal of four genes (Figure 4B).

**Figure 4.**
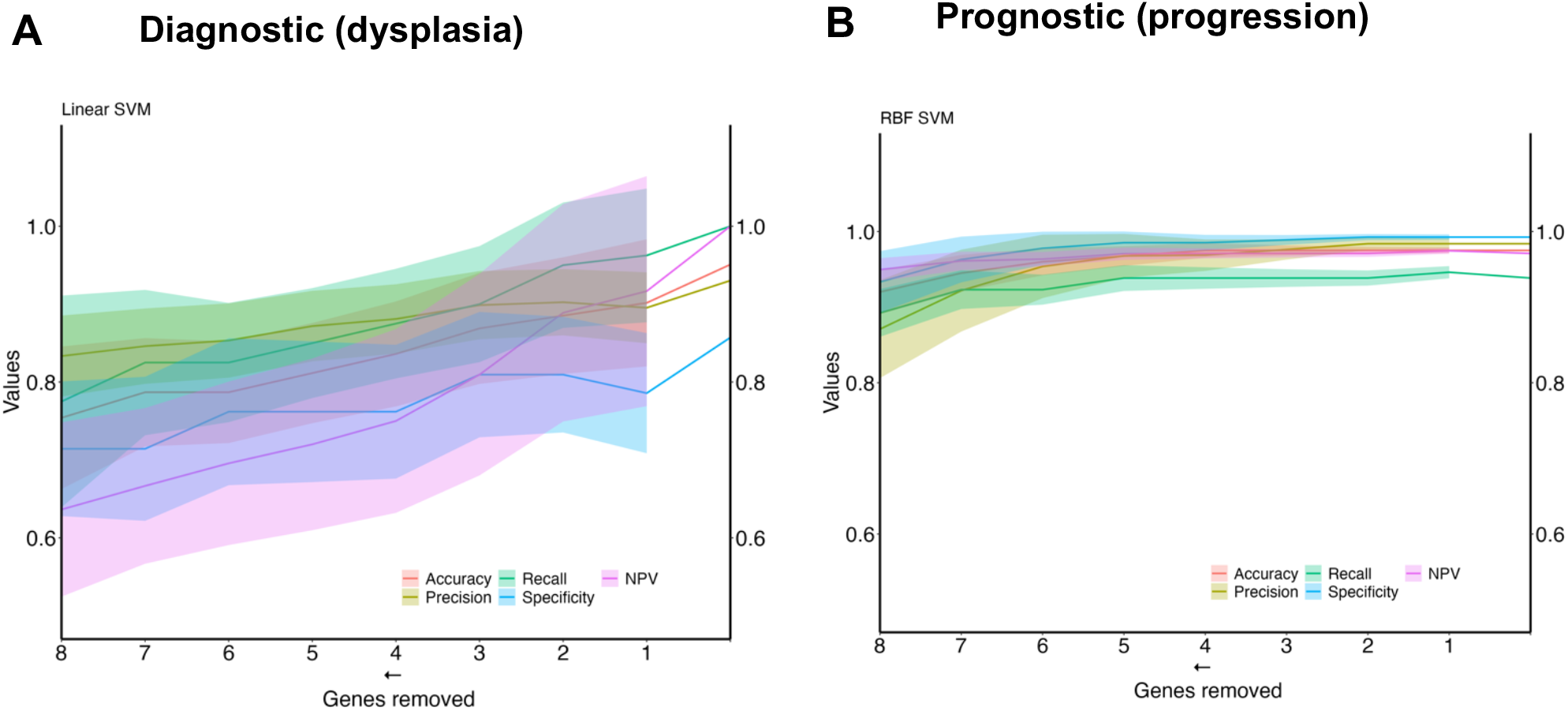
Metrics performance of the best algorithm in distinguishing ND-BE and LGD BE (diagnostic) and nonP-BE from P-BE (prognostic) when decreasing the number of genes included in the training. Mean of each performance metric (solid lines) and its respective standard deviation (ribbons) for the diagnostic algorithm, Linear Support Vector Machine (LSVM) (A) and for the prognostic algorithm, Radial Basis Function Support Vector Machine (RBF SVM) algorithm (B) NPV – negative predictive value.

### 3.5. *In-vivo* validation of key diagnostic and prognostic biomarkers

We conducted a validation study of the panel of biomarkers to distinguish between different stages of BE progression. Specifically, we performed RT-qPCR analysis to compare the expression levels of these biomarkers in differente cell lines: metaplasia (BAR-T and BAR-T10), dysplasia (CP-B, CP-C and CP-D), and EAC (OE33, KYAE-1 and ESO26). Each biomarker was tested with three technical replicates in each cell line.

For dysplasia diagnosis, we analyzed the expression of biomarkers in both metaplasia and dysplasia cell lines (Figure S6). In evaluating EAC prognosis, we compared the expression levels between metaplasia and EAC cell lines (Figure S7). Normalized expression values against reference genes (*PGK1*, *ELF1* and *RPL13A*) highlighted significant differences in key markers. For instance, biomarkers such as *IFI27* and *ATF3* differentiated metaplasia from dysplasia with statistically significant p-values (p = 0.009 and p = 0.003, respectively), revealing their potential utility in dysplasia diagnosis. Similarly, *CEBPB*, *SNAI1* and *CCN1* (alias *CYR61*) genes showed significant expression changes between metaplasia and EAC (p-values of 0,031, 0.022 and 0.038, respectively). This supports their relevance for EAC prediction. The observed differential expression patterns suggest that these biomarkers serve as valuable molecular tools for early detection of dysplasia and the risk of progression to EAC, facilitating timely clinical intervention. Interestingly, some of the top-performing genes, namely *IGHV3-43*, *IGHV4-3*1, *IGHV3-53*, and *PGC*, showed no detectable expression in cell lines. Since these genes ranked high according to the diagnostic algorithm, we hypothesized that their expression originates from immune cells, typically absent in cell lines. To investigate this further, we specifically tested these genes in tissue samples from BE patients with and without dysplasia. Contrary to cell lines, the expression of these genes was detectable in patient samples, supporting the notion that immune cells-may play a critical role in BE progression (Figure S6).

## 4. DISCUSSION

BE is the only known precursor to EAC, a malignancy with rising incidence and poor prognosis. This underscores the need for more effective management methods, including assertive early diagnosis of dysplasia and prognostic prediction within BE surveillance programs. While tools incorporating biomarkers are continuously emerging, few have reached clinical validation and implementation. Even fewer combine biomarkers with AI and those under clinical validation or use, do not provide simultaneous detection of dysplasia and prognostic assessment. Moreover, none can simultaneously achieve high sensitivity (recall) and high specificity.

In this study, we developed two algorithms to assist with the diagnosis of dysplasia, the prognosis of BE, and ultimately the management of EAC. The genes of interest for dysplasia detection (diagnostic algorithm) were newly identified from the raw data of three different RNAseq datasets. Conversely, the algorithm developed for prognosis was based on a gene set identified in a previous study [35]. For both applications, genes were ranked based on their F1 score, sensitivity (*aka* recall or true positive rate) and precision (*aka* positive predictive values) in predicting conditions such as LGD BE and P-BE.

In high-risk disease detection cases such as dysplasia, recall is a more important evaluation metric than precision because it can correctly identify all relevant positive cases (*i.e.*, samples containing dysplasia or at high risk of progressing to EAC). However, precision, which is the fraction of positive cases among all cases classified as positives by the model, is also crucial because it emphasizes the correctness of positive predictions made by the model (*i.e.*, measures how many cases are incorrectly classified as positive). In a situation where false positives have significant implications, such as subjecting BE patients without dysplasia or with a low risk of progression to unnecessary treatments or screening intervention, precision matters. Since both high precision and high recall were desirable for the present study, the ranking was based on the F1 score, which combines precision and recall using their harmonic mean. Maximizing the F1 score implies maximizing both precision and recall simultaneously. Performance metrics for each gene at its best threshold were high (see Supplementary Tables 5 and 6). However, specificity and NPV were higher for the prognostic genes, showing their great potential to exclude patients who are not at risk for malignant progression.

To better explore the potential predictive value of the selected biomarkers, we trained machine learning algorithms testing all possible combinations of biomarkers in each gene set. The average metrics of the newly trained algorithms with combinations of biomarkers showed increased predictive power (Table 2) compared to the predictive power of individual genes (Supplementary Tables 5 and 6), which is expected in the context of complex gene interactions. Finally, envisioning the clinical applicability of both algorithms, we evaluated the minimal number of biomarkers necessary to maintain high-performance metrics (LSVM for diagnosis and RBF SVM for prognosis) in each gene set. Both algorithms were tested with a decreasing number of genes, and as depicted in Figure 4, a reduction in performance metrics was observed when removing one gene from the diagnostic set and four genes from the prognostic’s gene set.

For diagnostic application, ten genes-*IGHV3-43*, *SLC38A4*, *PLLP*, *CELA3A*, *IGHV4-31*, *TMPRSS5*, *TP53*, *NR4A1*, *ATF3*, *IFI27*-were identified as the top candidates for dysplasia detection, particularly for distinguishing between NDBE and LGD BE. These genes are associated with different aspects of cancer biology, such as metabolism, cell invasion, and oncogenic processes, suggesting their potential as biomarkers in the context of BE dysplasia [61–64]. Moreover, transcription factors such as *NR4A1* and *ATF3*, have been previously associated with BE with LGD [65].

For the prognostic application, an RBF SVM algorithm was selected, which uses the expression pattern of ten genes (*SNAI1*, *C1S*, *DUSP1*, *CEBPB*, *COL4A1*, *ZEB1*, *CEBPD*, *CCN1*, *LAMC1* and *TWIST1*). Four of these genes – *SNAI1, COL4A1, ZEB1,* and *TWIST* – have been associated with epithelial-to-mesenchymal transition [66]. *COL4A1*, *ZEB1,* and *TWIST1* have also been described as potential screening biomarkers of BE malignant progression. *COL4A1* is upregulated in EAC versus BE [67–69] and is associated with poor EAC prognosis [68], and it predicts the response to immune checkpoint inhibitors in EAC [67]. Increased expression of *ZEB1* has been associated with the repression of *CDH1* [70], which is associated with BE progression to EAC [71–75]. *TWIST1* up-regulation was observed in at-risk BE samples years before the emergence of any microscopic signs of malignancy (dysplasia/EAC) [35].

The genes *TP53* and *CDH1* were included in both gene sets to train the classifiers. *TP53* is known for its role in BE malignant progression [76, 77], improved prediction of BE neoplastic progression [78], increased risk of dysplasia when abnormally expressed, and improved intra-observer agreement in dysplastic diagnosis [79]. *CDH1* has severely reduced or disorganized expression during BE dysplastic progression [reviewed by [80] and an almost undetectable expression in poorly differentiated EAC [71–75]. While *TP53* alone is insufficient for diagnostic and for prognostic applications, it has been shown to have predictive value in combination with other biomarkers in the diagnostic setting. These findings confirm the previously studied role of *TP53* in the pathogenesis of BE dysplasia [81, 82]. Because *TP53* mutations are often associated with a higher risk of progression in BE patients [83], further validation of this biomarker at the molecular level, including its mutational status and RNA expression levels, is warranted.

All metrics of both algorithms are higher when compared to currently available tools for risk stratification, such as *TP53* immunohistochemistry (0.49 recall/sensitivity, 0.86 specificity [81]) and TissueCypher (0.55 recall/sensitivity, 0.82 specificity for high-intermediate risk class 55%/82%) [63]. Tools for dysplasia detection, such as Wats3D and Cytosponge-*TFF3*. are still under prospective evaluation. Wats3D provides an incremental yield of 7% for any dysplasia subtype but is negative for dysplasia in 62.5% of cases where an endoscopic biopsy confirmation to compare with the gold standard revealed dysplasia [86]. The Cytosponge-*TFF3* test when combined with a multidimensional biomarker panel and fitted into a regression model was shown to be able to predict patients with dysplasia with good accuracy but further validation is still needed [87]. Interestingly, in our top 45 genes for diagnostic application (Supplementary Table 4), we have identified another trefoil factor, the *TFF2*, which is BE related gene.

A preliminary *in vivo* validation of the selected diagnostic and prognostic biomarkers was conducted by examining their expression in metaplasia, dysplasia and EAC-derived cell lines. This validation confirmed their differential expression, highlighting their potential in distinguishing BE progression stages. Exceptionally, *IGHV3-43*, *IGHV4-31*, *IGHV3-53*, and *PGC* top-ranked genes were validated in FFPE samples from patients diagnosed with BE with and without dysplasia due to their lack of expression in the cell lines. The absence of immune cells in cell line cultures, which focus on epithelial cells, likely contributes to these findings. While we cannot exclude that the used cell lines may exhibit genetic differences from the original tissue, which potentially influences their molecular profiles [88], further clinical validation with a selected cohort of patient samples is warranted and is currently underway.

No molecular tools are currently implemented in clinical practice for identifying LGD/HGD BE. Dysplasia is a major biomarker in BE risk stratification, but it is often focal, making accurate characterization of collected BE biopsy challenging [89], and leading to many cases of BE classified as INDBE. INDBE is a management limbo for dysplasia, posing problems for clinicians.

Most clinical tools developed for BE focus on risk stratification (prognosis) [28, 37–39, 84, 85, 90] and have a high specificity (identify and correctly exclude BE patients not at risk of progression). Simultaneously, these tools have a lower recall/sensitivity indicating their performance drops in detecting BE patients at true risk of progression.

New tests that aim for high recall and sensitivity are vital to avoid missing unacceptable true positive cases of LGD or HGD, as well as patients at risk of progression. However, these tests must also maintain high precision and high sensitivity to avoid incorrectly including patients not having dysplasia or having a low risk of progression. This balance can improve surveillance of high-risk patients while reducing unnecessary procedures for low-risk patients, ultimately lowering patient management costs. Our approach, which combines machine learning algorithms with gene expression signatures, represents a promising breakthrough in healthcare. It has the potential to significantly enhance both the diagnosis and prognosis of dysplasia by delivering high recall and precision into clinical practice.

## 5. CONCLUSIONS

This study not only identified biomarkers and developed algorithms to detect LGD in BE biopsies and predict the progression of BE to EAC, but also paved the way for creating new *in-vitro* laboratory tests for the diagnosis and prognosis of BE. Both algorithms were developed using datasets from public databases analyzing tissue samples obtained during routine endoscopy.

For the prediction of BE malignant progression, an LSVM algorithm featuring the identification of LGD was trained while an RBF SVM algorithm was trained for the prediction of BE malignant progression. Both algorithms reached high-performance metrics. To our knowledge, no existing tools can simultaneously detect dysplasia and assess the risk of progression with such high precision and recall.

Validation of the biomarkers and algorithms presented in this study in an independent test and validation patient cohort is currently under consideration. Additionally, while no other known risk factors (epidemiologic, clinical, histologic) have been combined with the presented biomarkers, incorporating patient demographic and clinical information could further enhance the predictive value of the gene expression algorithms. Future algorithm developments will address this issue, demonstrating how such combinations can significantly boost their predictive power.

## Data Availability

All data produced in the present study are available upon reasonable request to the authors

https://www.ncbi.nlm.nih.gov/geo/

## AUTHOR CONTRIBUTIONS

Migla Miskinyte: Conceptualization, Formal analysis, Investigation, Writing – review & editing. Benilde Pondeca: Conducting experiments, Writing – review & editing. José B. Pereira-Leal: Conceptualization, review & editing. Joana Cardoso: Conceptualization, Validation, Investigation, Writing – review & editing. All authors have read and agreed to the published version of the manuscript.

## Supplementary Material

Supplementary Table 1 List of genes used for differential expression analysis across multiple datasets

Supplementary Table 2 List of differentially expressed genes between NDBE and LGDBE groups, E_MTAB_4054 dataset.

Supplementary Table 3 List of differentially expressed genes between NDBE and LGDBE groups, GSE58963dataset.

Supplementary Table 4 Top 45 candidate genes.

**Supplementary Table 5.** Top 15 genes for prognostic: individual genes’ predictive metrics.

| N | Gene | Threshold | F1-score | Recall | Precision | Specificity | NPV | Accuracy | FPR | TP | FP | TN | FN |
| --- | --- | --- | --- | --- | --- | --- | --- | --- | --- | --- | --- | --- | --- |
| 1 | *COL4A1* | 9.34 | 0.84 | 0.75 | 0.94 | 0.98 | 0.89 | 0.91 | 0.02 | 49 | 3 | 132 | 16 |
| 2 | *LAMC1* | 9.19 | 0.84 | 0.88 | 0.81 | 0.90 | 0.94 | 0.90 | 0.10 | 57 | 13 | 122 | 8 |
| 3 | *CEBPB* | 9.75 | 0.84 | 0.80 | 0.88 | 0.95 | 0.91 | 0.90 | 0.05 | 52 | 7 | 128 | 3 |
| 4 | *CCN1* | 7.68 | 0.84 | 0.88 | 0.80 | 0.90 | 0.94 | 0.89 | 0.10 | 57 | 14 | 121 | 8 |
| 5 | *SNAI1* | 6.61 | 0.82 | 0.97 | 0.71 | 0.81 | 0.98 | 0.86 | 0.19 | 63 | 26 | 109 | 2 |
| 6 | *C1S* | 9.65 | 0.83 | 0.83 | 0.83 | 0.92 | 0.92 | 0.89 | 0.08 | 54 | 11 | 124 | 11 |
| 7 | *ZEB1* | 7.66 | 0.80 | 0.91 | 0.72 | 0.83 | 0.95 | 0.86 | 0.17 | 59 | 23 | 112 | 6 |
| 8 | *CEBPD* | 9.32 | 0.81 | 0.83 | 0.79 | 0.90 | 0.92 | 0.88 | 0.10 | 54 | 14 | 121 | 11 |
| 9 | *DUSP1* | 10.46 | 0.81 | 0.82 | 0.80 | 0.90 | 0.91 | 0.88 | 0.10 | 53 | 13 | 122 | 12 |
| 10 | *VWF* | 8.88 | 0.77 | 0.80 | 0.74 | 0.87 | 0.90 | 0.85 | 0.13 | 52 | 18 | 117 | 13 |
| 11 | *TWIST* | 5.15 | 0.67 | 1.00 | 0.51 | 0.53 | 1.00 | 0.69 | 0.47 | 65 | 63 | 72 | 0 |
| 12 | *PLPP3* | 8.68 | 0.68 | 0.94 | 0.53 | 0.60 | 0.95 | 0.71 | 0.40 | 61 | 54 | 81 | 4 |
| 13 | *ACTN1* | 9.82 | 0.73 | 0.74 | 0.72 | 0.86 | 0.87 | 0.82 | 0.14 | 48 | 19 | 116 | 17 |
| 14 | *CDH1* | 5.70 | 0.50 | 1.00 | 0.33 |  |  | 0.33 | 1.00 | 65 | 135 | 0 | 0 |
| 15 | *TP53* | 7.12 | 0.62 | 0.89 | 0.48 | 0.53 | 0.91 | 0.65 | 0.47 | 58 | 64 | 71 | 7 |

**Supplementary Table 6.** Top 18 genes for diagnostic: individual genes’ predictive metrics.

| N | Gene | Threshold | F1-score | Recall | Precision | Specificity | NPV | Accuracy | FPR | TP | FP | TN | FN |
| --- | --- | --- | --- | --- | --- | --- | --- | --- | --- | --- | --- | --- | --- |
| 1 | *SLC38A4* | -0.21 | 0.84 | 0.90 | 0.78 | 0.52 | 0.73 | 0.77 | 0.48 | 36 | 10 | 11 | 4 |
| 2 | *TMPRSS5* | -1.14 | 0.82 | 0.98 | 0.71 | 0.24 | 0.83 | 0.72 | 0.76 | 39 | 16 | 5 | 1 |
| 3 | *EGR3* | 0.04 | 0.82 | 0.98 | 0.71 | 0.24 | 0.83 | 0.72 | 0.76 | 39 | 16 | 5 | 1 |
| 4 | *TP53* | 5.05 | 0.82 | 0.95 | 0.72 | 0.29 | 0.75 | 0.72 | 0.71 | 38 | 15 | 6 | 2 |
| 5 | *FOSB* | 1.15 | 0.82 | 1.00 | 0.69 | 0.14 | 1.00 | 0.70 | 0.86 | 40 | 18 | 3 | 0 |
| 6 | *NR4A1* | 3.22 | 0.81 | 1.00 | 0.68 | 0.10 | 1.00 | 0.69 | 0.90 | 40 | 19 | 2 | 0 |
| 7 | *SFTPB* | -0.10 | 0.80 | 0.95 | 0.69 | 0.19 | 0.67 | 0.69 | 0.81 | 38 | 17 | 4 | 2 |
| 8 | *IFI27* | 3.87 | 0.79 | 0.98 | 0.66 | 0.05 | 0.50 | 0.66 | 0.95 | 39 | 20 | 1 | 1 |
| 9 | *PLLP* | 5.66 | 0.78 | 0.95 | 0.66 | 0.05 | 0.33 | 0.64 | 0.95 | 38 | 20 | 1 | 2 |
| 10 | *CELA3A* | -3.84 | 0.78 | 0.95 | 0.66 | 0.05 | 0.33 | 0.64 | 0.95 | 38 | 20 | 1 | 2 |
| 11 | *ATF3* | 2.94 | 0.78 | 0.95 | 0.66 | 0.05 | 0.33 | 0.64 | 0.95 | 38 | 20 | 1 | 2 |
| 12 | *IGHV3-43* | -1.38 | 0.78 | 0.95 | 0.66 | 0.05 | 0.33 | 0.64 | 0.95 | 38 | 20 | 1 | 2 |
| 13 | *CDH1* | 8.61 | 0.78 | 0.95 | 0.66 | 0.05 | 0.33 | 0.64 | 0.95 | 38 | 20 | 1 | 2 |
| 14 | *PGC* | 2.48 | 0.74 | 0.88 | 0.64 | 0.05 | 0.17 | 0.59 | 0.95 | 35 | 20 | 1 | 5 |
| 15 | *GKN2* | -0.11 | 0.74 | 0.88 | 0.64 | 0.05 | 0.17 | 0.59 | 0.95 | 35 | 20 | 1 | 5 |
| 16 | *IGHV4-31* | -0.91 | 0.74 | 0.88 | 0.64 | 0.05 | 0.17 | 0.59 | 0.95 | 35 | 20 | 1 | 5 |
| 17 | *IGHV3-53* | -0.92 | 0.74 | 0.88 | 0.64 | 0.05 | 0.17 | 0.59 | 0.95 | 35 | 20 | 1 | 5 |
| 18 | *PNLIPRP1* | -2.73 | 0.72 | 0.83 | 0.63 | 0.10 | 0.22 | 0.57 | 0.90 | 33 | 19 | 2 | 7 |

**Supplementary Table 7.** Gene frequency in the best-performing algorithms for diagnostic (LSVM = 1115)

| Gene | Count | Frequency (%) |
| --- | --- | --- |
| *IGHV3-43* | 1115 | 100.00 |
| *SLC38A4* | 1097 | 98.39 |
| *PLLP* | 1072 | 96.14 |
| *CELA3A* | 923 | 82.78 |
| *IGHV4-31* | 850 | 76.23 |
| *TMPRSS5* | 646 | 57.94 |
| *TP53* | 645 | 57.85 |
| *NR4A1* | 633 | 56.77 |
| *ATF3* | 625 | 56.05 |
| *IFI27* | 581 | 52.11 |
| *PGC* | 530 | 47.53 |
| *GKN2* | 490 | 43.95 |
| *PNLIPRP1* | 470 | 42.15 |
| *SFTPB* | 460 | 41.26 |
| *CDH1* | 379 | 34.00 |
| *FOSB* | 369 | 33.10 |
| *EGR3* | 355 | 31.84 |
| *IGV3-53* | 330 | 29.60 |

**Supplementary Table 8.** Gene frequency in the best-performing algorithms for prognostic (RBF SVM = 5794)

| Gene | Count | Frequency (%) |
| --- | --- | --- |
| *SNAI1* | 5545 | 95.70245 |
| *DUSP1* | 3837 | 66.22368 |
| *CEBPB* | 3791 | 65.42975 |
| *C1S* | 3672 | 63.37591 |
| *COL4A1* | 3409 | 58.83673 |
| *LAMC1* | 3157 | 54.4874 |
| *CEBPD* | 3121 | 53.86607 |
| *ZEB1* | 3105 | 53.58992 |
| *TWIST1* | 2940 | 50.74215 |
| *CCN1* | 2913 | 50.27615 |
| *TP53* | 2872 | 49.56852 |
| *ACTN1* | 2609 | 45.02934 |
| *VWF* | 2592 | 44.73593 |
| *PLPP3* | 2336 | 40.31757 |
| *CDH1* | 1140 | 19.67553 |

**Figure S1.**
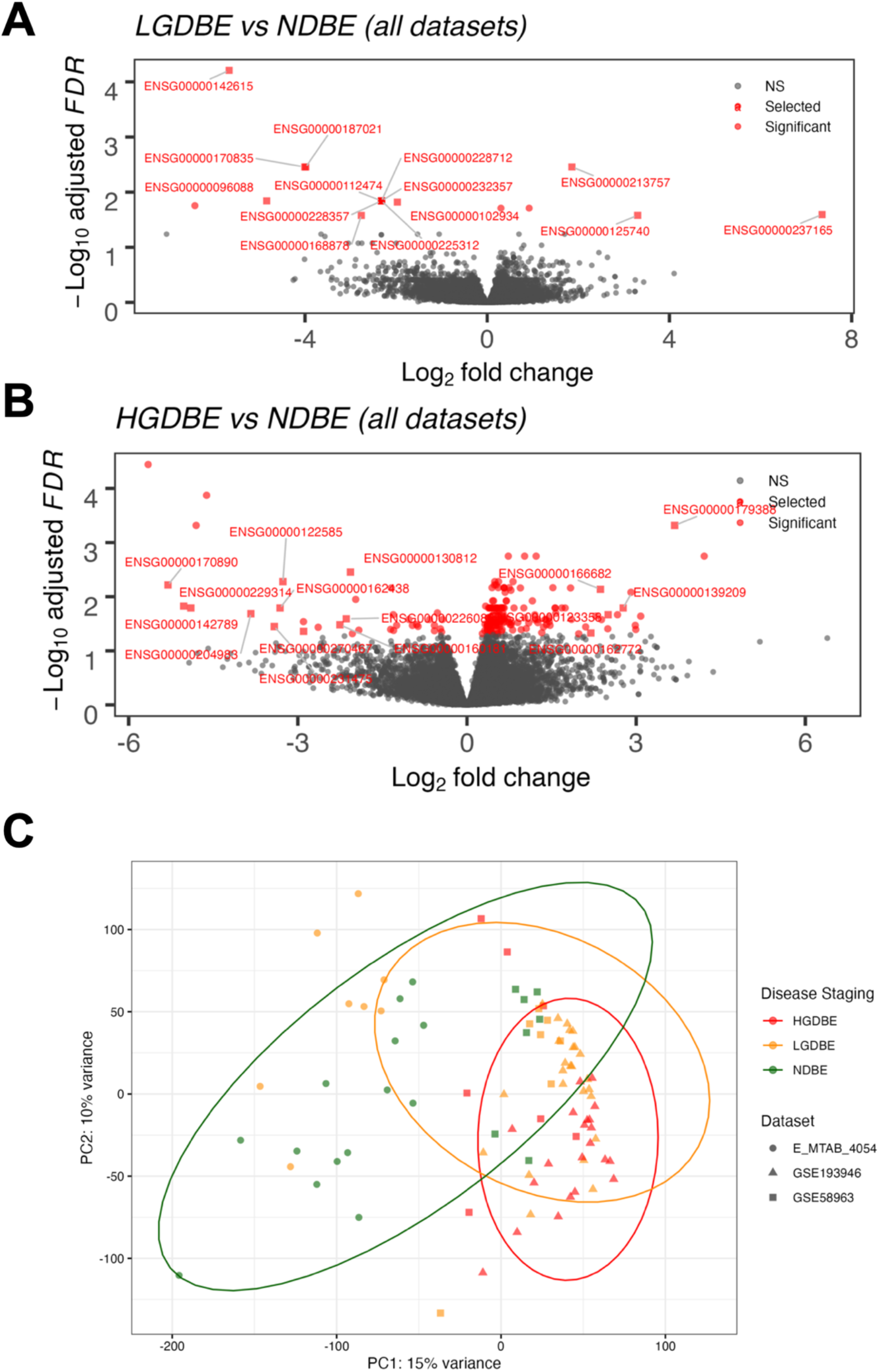
Identification of 30 biomarkers using all datasets.

**Figure S2.**
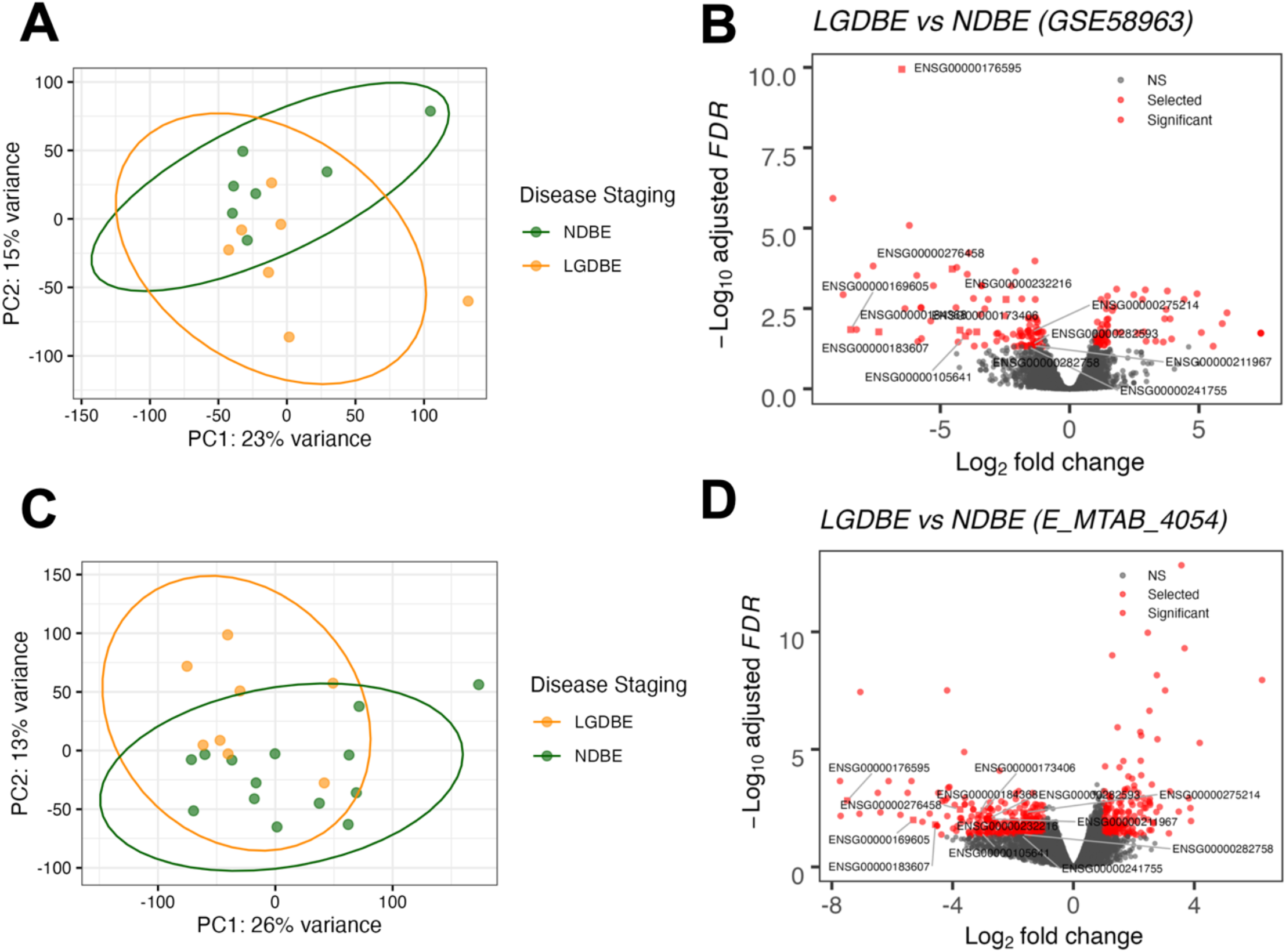
Identification of additional 13 biomarkers by analyzing datasets separately.

**Figure S3.**
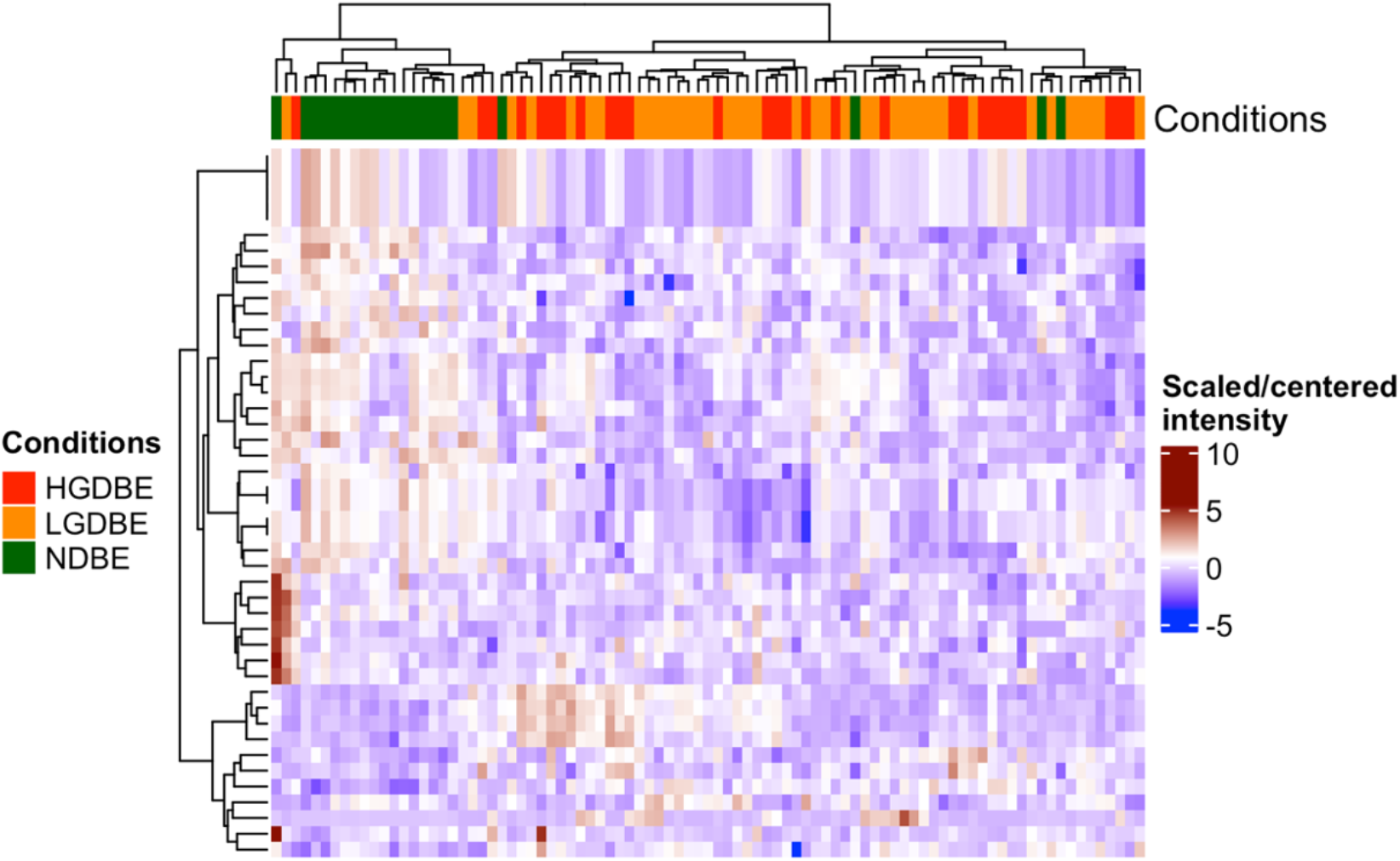
Heatmap of top 45 genes selected for diagnostics.

**Figure S4.**
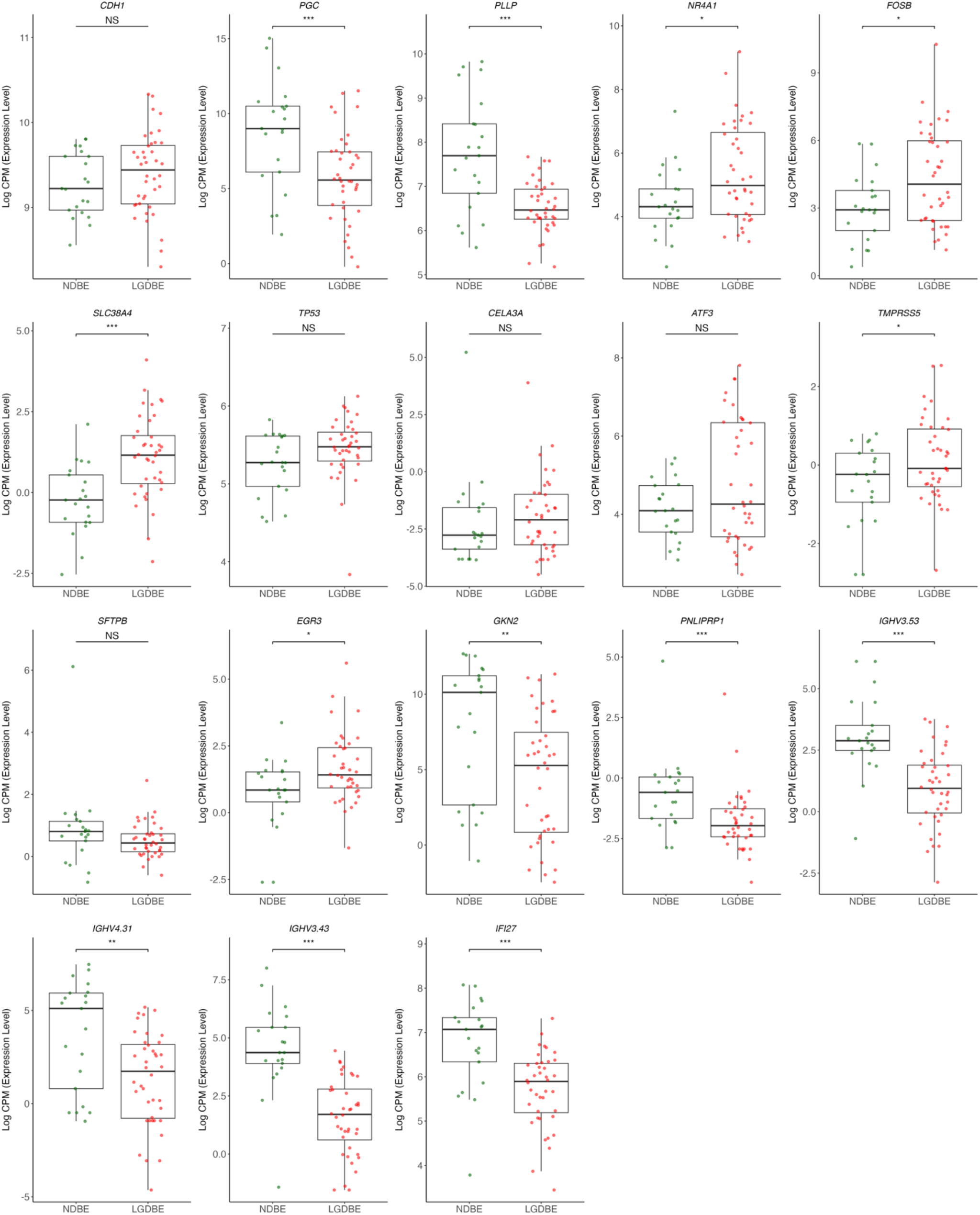
Boxplots of gene expression levels of genes potentially associated with low-grade dysplasia. Comparison of each gene expression in NDBE and LGDBE samples obtained from a total of 3 datasets. *** adj. *p <* 0.001; ** adj. *p <* 0.01; * adj. *p <* 0.05; ^NS^ adj. *p* > 0.05.

**Figure S5.**
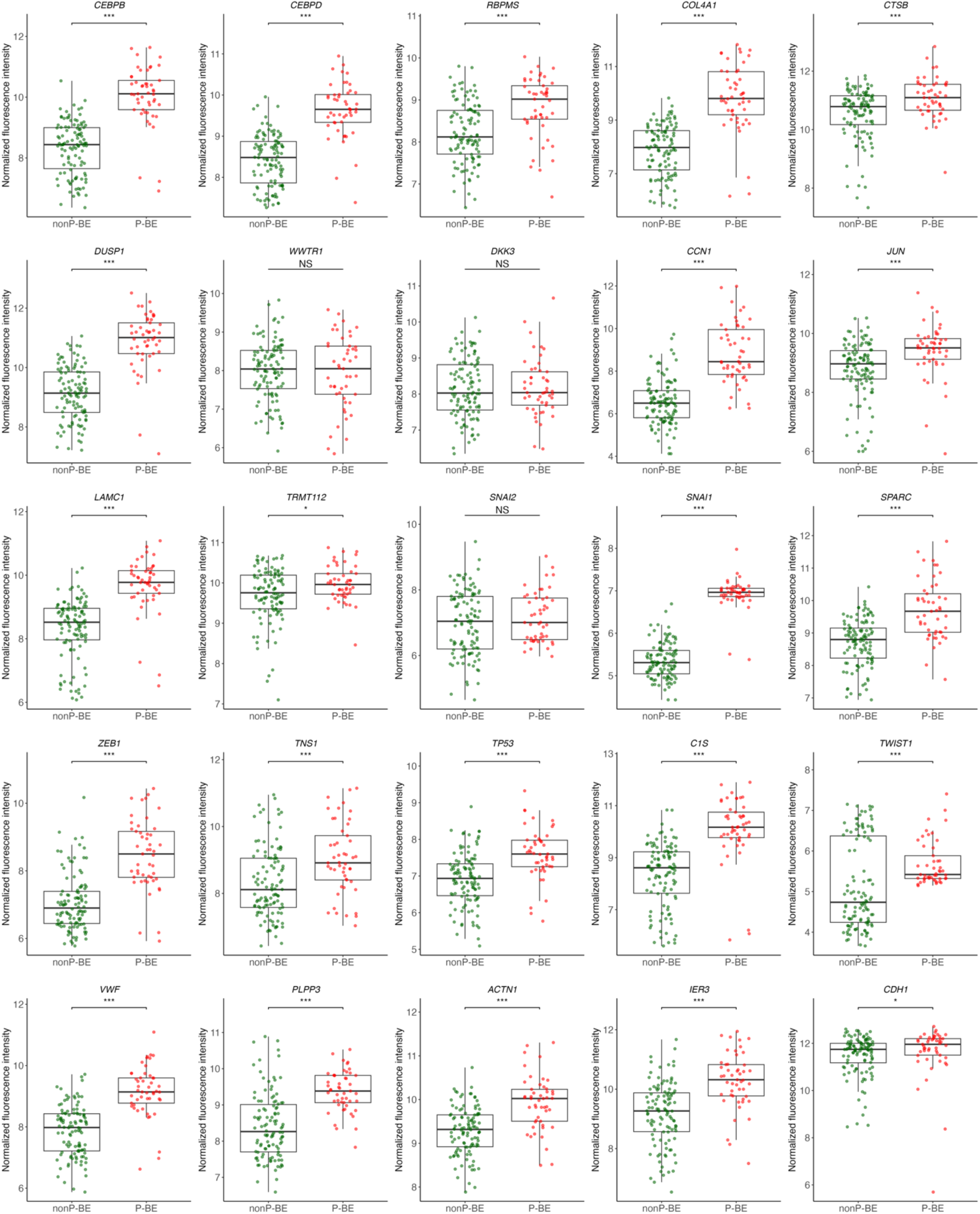
Boxplots of gene expression levels of genes potentially associated with prognosis. Comparison of each gene expression in nonP-BE and P-BE samples obtained from a total of 9 datasets. *** adj. p < 0.001; ** adj. p < 0.01; * adj. p < 0.05; NS adj. p > 0.05.

**Figure S6.**
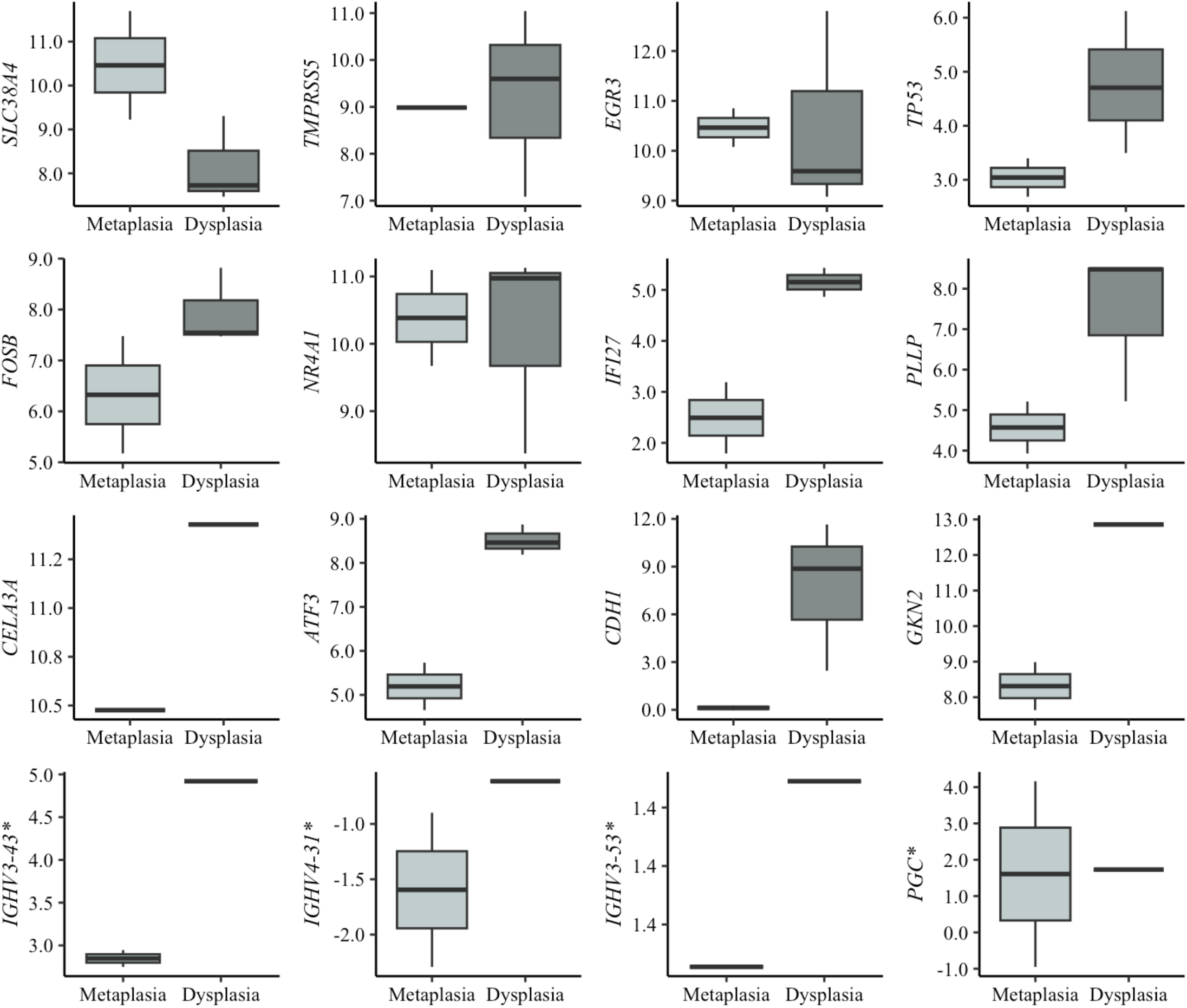
Boxplots of gene expression levels of diagnostic genes. Comparison of gene expression levels between cell lines representing metaplasia and dysplasia, with expression levels normalized to reference genes (*PGK1, ELF1*, and *RPL13A*). * Genes were additionally tested in FFPE samples from patients diagnosed with BE with and without dysplasia.

**Figure S7.**
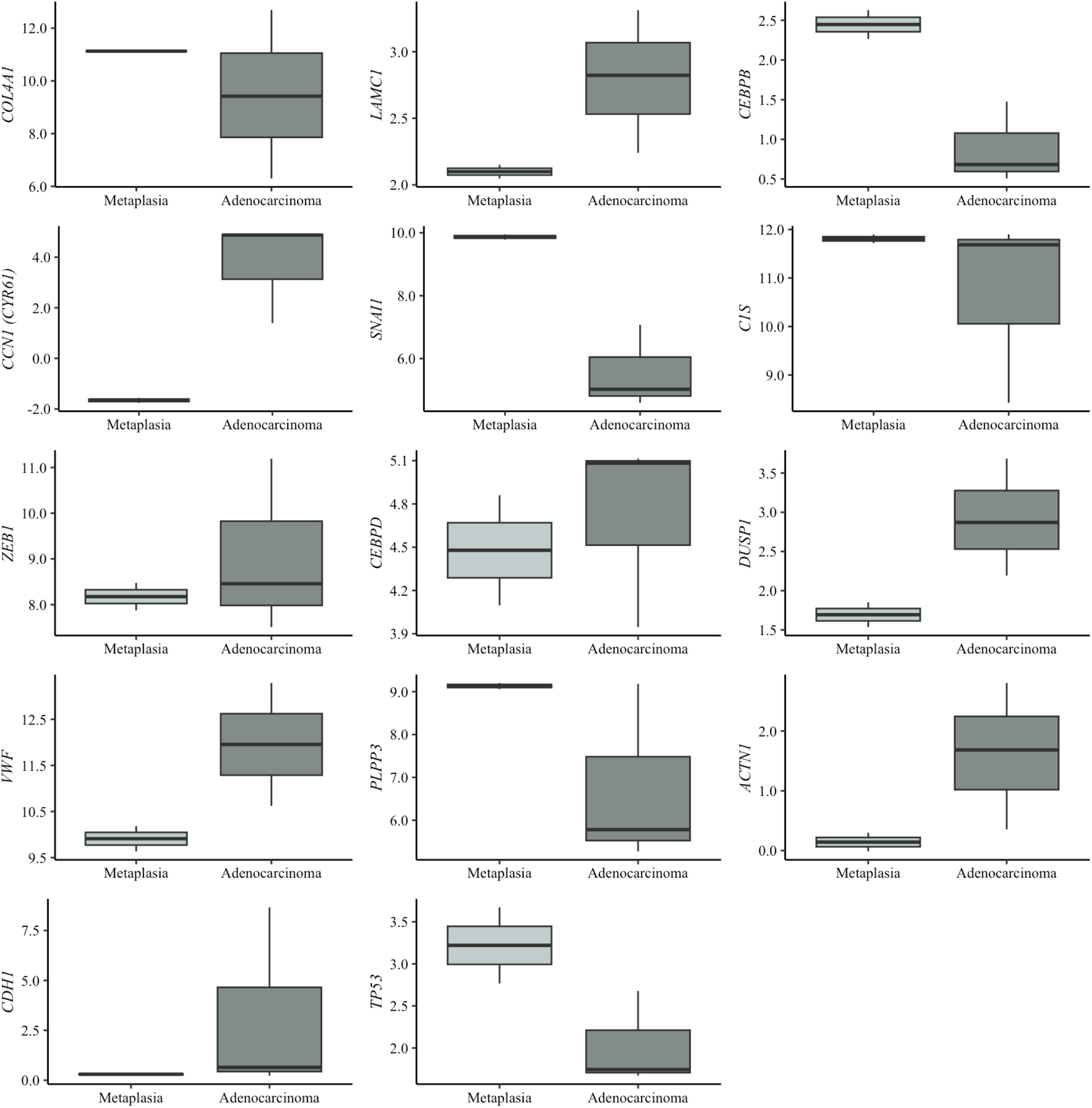
Boxplots of gene expression levels of prognostic genes. Comparison of gene expression levels between cell lines representing metaplasia and EAC, with expression levels normalized to reference genes (*PGK1*, *ELF1*, and *RPL13A*).

## REFERENCES

1. Spechler SJ, Souza RF. Barrett’s Esophagus. New England Journal of Medicine. 2014;371:836–45.

2. Choi KKH, Sanagapalli S. Barrett’s esophagus: Review of natural history and comparative efficacy of endoscopic and surgical therapies. World J Gastrointest Oncol. 2022;14:568–86.

3. Klavan H, Russell MB, Macklin J, Lee E, Aslanian HR, Muniraj T. Barrett’s esophagus: A comprehensive review for the internist. Disease-a-Month. 2018;64:471–87.

4. Killcoyne S, Fitzgerald RC. Evolution and progression of Barrett’s oesophagus to oesophageal cancer. Nature Reviews Cancer. 2021;21:731–41.

5. Malfertheiner P, Nocon M, Vieth M, Stolte M, Jaspersen D, Koelz HR, et al. Evolution of gastro-oesophageal reflux disease over 5 years under routine medical care--the ProGERD study. Aliment Pharmacol Ther. 2012;35:154–64.

6. Fabian T, Leung A. Epidemiology of Barrett’s Esophagus and Esophageal Carcinoma. Surgical Clinics of North America. 2021;101:381–9.

7. Cook MB, Shaheen NJ, Anderson LA, Giffen C, Chow WH, Vaughan TL, et al. Cigarette smoking increases risk of Barrett’s esophagus: An analysis of the barrett’s and esophageal adenocarcinoma consortium. Gastroenterology. 2012;142:744–53.

8. Sinha DN, Abdulkader RS, Gupta PC. Smokeless tobacco-associated cancers: A systematic review and meta-analysis of Indian studies. Int J Cancer. 2016;138:1368–79.

9. Kamat P, Wen S, Morris J, Anandasabapathy S. Exploring the Association Between Elevated Body Mass Index and Barrett’s Esophagus: A Systematic Review and Meta-Analysis. Annals of Thoracic Surgery. 2009;87:655–62.

10. Andrici J, Tio M, Cox MR, Eslick GD. Hiatal hernia and the risk of Barrett’s esophagus. Journal of Gastroenterology and Hepatology (Australia). 2013;28:415–31.

11. Mittal SK, Abdo J, Adrien MP, Bayu BA, Kline JR, Sullivan MM, et al. Current state of prognostication, therapy and prospective innovations for Barrett’s-related esophageal adenocarcinoma: a literature review. J Gastrointest Oncol. 2021;12:1197–214.

12. Sung H, Ferlay J, Siegel RL, Laversanne M, Soerjomataram I, Jemal A, et al. Global Cancer Statistics 2020: GLOBOCAN Estimates of Incidence and Mortality Worldwide for 36 Cancers in 185 Countries. CA Cancer J Clin. 2021;71:209–49.

13. Morgan E, Soerjomataram I, Rumgay H, Coleman HG, Thrift AP, Vignat J, et al. The Global Landscape of Esophageal Squamous Cell Carcinoma and Esophageal Adenocarcinoma Incidence and Mortality in 2020 and Projections to 2040: New Estimates From GLOBOCAN 2020. Gastroenterology. 2022;163:649–658.e2.

14. Then EO, Lopez M, Saleem S, Gayam V, Sunkara T, Culliford A, et al. Esophageal cancer: An updated surveillance epidemiology and end results database analysis. World J Oncol. 2020;11:55–64.

15. Fabian T, Leung A. Epidemiology of Barrett’s Esophagus and Esophageal Carcinoma. Surgical Clinics of North America. 2021;101:381–9.

16. Hamade N, Vennelaganti S, Parasa S, Vennalaganti P, Gaddam S, Spaander MCW, et al. Lower Annual Rate of Progression of Short-Segment vs Long-Segment Barrett’s Esophagus to Esophageal Adenocarcinoma. Clin Gastroenterol Hepatol. 2019;17:864.

17. Li N, Yang WL, Cai MH, Chen X, Zhao R, Li MT, et al. Burden of gastroesophageal reflux disease in 204 countries and territories, 1990–2019: a systematic analysis for the Global Burden of disease study 2019. BMC Public Health. 2023;23:1–13.

18. Zhang D, Liu S, Li Z, Wang R. Global, regional and national burden of gastroesophageal reflux disease, 1990–2019: update from the GBD 2019 study. Ann Med. 2022;54:1372–84.

19. Spechler SJ, Sharma P, Souza RF, Inadomi JM, Shaheen NJ. American gastroenterological association technical review on the management of Barrett’s esophagus. Gastroenterology. 2011;140.

20. Lee SW, Lien HC, Chang C Sen, Lin MX, Chang CH, Ko CW. Benefits of the Seattle biopsy protocol in the diagnosis of Barrett’s esophagus in a Chinese population. World J Clin Cases. 2018;6:753–8.

21. Eluri S, Shaheen NJ. Barrett’s Esophagus: Diagnosis and Management. Gastrointest Endosc. 2017;85:889.

22. Visrodia K, Singh S, Krishnamoorthi R, Ahlquist DA, Wang KK, Iyer PG, et al. Magnitude of Missed Esophageal Adenocarcinoma After Barrett’s Esophagus Diagnosis: A Systematic Review and Meta-analysis. Gastroenterology. 2016;150:599–607.e7.

23. Sharma P. Low-grade dysplasia in Barrett’s esophagus. Gastroenterology. 2004;127:1233– 8.

24. Runge TM, Abrams JA, Shaheen NJ. Epidemiology of Barrett’s Esophagus and Esophageal Adenocarcinoma. Gastroenterology Clinics of North America. 2015;44:203–31.

25. Alshelleh M, Inamdar S, McKinley M, Stewart M, Novak JS, Greenberg RE, et al. Incremental yield of dysplasia detection in Barrett’s esophagus using volumetric laser endomicroscopy with and without laser marking compared with a standardized random biopsy protocol. Gastrointest Endosc. 2018;88:35–42.

26. Thota PN, Kistangari G, Esnakula AK, Gonzalo DH, Liu X-L. Clinical significance and management of Barrett’s esophagus with epithelial changes indefinite for dysplasia. World J Gastrointest Pharmacol Ther. 2016;7:406–11.

27. Shaheen NJ, Falk GW, Iyer PG, Souza RF, Yadlapati RH, Sauer BG, et al. Diagnosis and Management of Barrett’s Esophagus: An Updated ACG Guideline. Am J Gastroenterol. 2022;117:559–87.

28. Vaughan TL, Onstad L, Dai JY. Interactive decision support for esophageal adenocarcinoma screening and surveillance. BMC Gastroenterol. 2019;19.

29. Galipeau PC, Li X, Blount PL, Maley CC, Sanchez CA, Odze RD, et al. NSAIDs Modulate CDKN2A, TP53, and DNA Content Risk for Progression to Esophageal Adenocarcinoma. PLoS Med. 2007;4:e67.

30. Trindade AJ, McKinley MJ, Alshelleh M, Levi G, Stewart M, Quinn KJ, et al. Mutational load may predict risk of progression in patients with Barrett’s oesophagus and indefinite for dysplasia: A pilot study. BMJ Open Gastroenterol. 2019;6.

31. Maley CC, Galipeau PC, Li X, Sanchez CA, Paulson TG, Blount PL, et al. The Combination of Genetic Instability and Clonal Expansion Predicts Progression to Esophageal Adenocarcinoma. 2004.

32. Mokrowiecka A, Wierzchniewska-ŁAwska A, Smolarz B, Romanowicz-Makowska H, Małecka-Panas E. P16 gene mutations in Barrett’s esophagus in gastric metaplasia - Intestinal metaplasia - Dysplasia - Adenocarcinoma sequence. Adv Med Sci. 2012;57:71–6.

33. Paulson TG, Maley CC, Li X, Li H, Sanchez CA, Chao DL, et al. Chromosomal instability and copy number alterations in Barrett’s esophagus and esophageal adenocarcinoma. Clinical Cancer Research. 2009;15:3305–14.

34. Merlo LMF, Shah NA, Li X, Blount PL, Vaughan TL, Reid BJ, et al. A comprehensive survey of clonal diversity measures in Barrett’s esophagus as biomarkers of progression to esophageal adenocarcinoma. Cancer Prevention Research. 2010;3:1388–97.

35. Cardoso J, Mesquita M, Dias Pereira A, Bettencourt-Dias M, Chaves P, Pereira-Leal JB. CYR61 and TAZ upregulation and focal epithelial to mesenchymal transition may be early predictors of barrett’s esophagus malignant progression. PLoS One. 2016;11.

36. Selaru FM, Zou T, Xu Y, Shustova V, Yin J, Mori Y, et al. Global gene expression profiling in Barrett’s esophagus and esophageal cancer: a comparative analysis using cDNA microarrays. Oncogene. 2022;21:475–8.

37. Moinova HR, Laframboise T, Lutterbaugh JD, Chandar AK, Dumot J, Faulx A, et al. Identifying DNA methylation biomarkers for non-endoscopic detection of Barrett’s esophagus. 2018.

38. Jin Z, Cheng Y, Gu W, Zheng Y, Sato F, Mori Y, et al. A multicenter, double-blinded validation study of methylation biomarkers for progression prediction in Barrett’s esophagus. Cancer Res. 2009;69:4112–5.

39. Abdo J, Wichman CS, Dietz NE, Ciborowski P, Fleegel J, Mittal SK, et al. Discovery of novel and clinically relevant markers in formalin-fixed paraffin-embedded esophageal cancer specimen. Front Oncol. 2018;8 MAY.

40. Tan JL, Chinnaratha MA, Woodman R, Martin R, Chen HT, Carneiro G, et al. Diagnostic Accuracy of Artificial Intelligence (AI) to Detect Early Neoplasia in Barrett’s Esophagus: A Non-comparative Systematic Review and Meta-Analysis. Front Med (Lausanne). 2022;9:890720.

41. Fouad YM, Mostafa I, Yehia R, EL-Khayat H. Biomarkers of Barrett’s esophagus. World J Gastrointest Pathophysiol. 2014;5:450.

42. Honing J, Fitzgerald RC. Categorizing Risks within Barrett’s Esophagus To Guide Surveillance and Interception; Suggesting a New Framework. Cancer Prev Res (Phila). 2023;16:313–20.

43. Mejza M, Małecka-Wojciesko E. Diagnosis and Management of Barrett’s Esophagus. J Clin Med. 2023;12.

44. Robinson MD, McCarthy DJ, Smyth GK. edgeR: A Bioconductor package for differential expression analysis of digital gene expression data. Bioinformatics. 2009;26:139–40.

45. Leek JT, Johnson WE, Parker HS, Jaffe AE, Storey JD. The sva package for removing batch effects and other unwanted variation in high-throughput experiments. Bioinformatics. 2012;28:882.

46. Watts GS, Tran NL, Berens ME, Bhattacharyya AK, Nelson MA, Montgomery EA, et al. Identification of Fn14/TWEAK receptor as a potential therapeutic target in esophageal adenocarcinoma. Int J Cancer. 2007;121:2132–9.

47. Jabeen A, Ahmad N, Raza K. Machine Learning-based state-of-the-art methods for the classification of RNA-Seq data. In: Classification in Bioapps: Automation of Decision Making. Springer; 2018. p. 133–72.

48. Pirooznia M, Yang JY, Qu MQ, Deng Y. A comparative study of different machine learning methods on microarray gene expression data. BMC Genomics. 2008;9 SUPPL. 1.

49. Peixoto C, Lopes MB, Martins M, Casimiro S, Sobral D, Grosso AR, et al. Identification of biomarkers predictive of metastasis development in early-stage colorectal cancer using network-based regularization. BMC Bioinformatics. 2023;24.

50. MacCarthy FP, Duggan SP, Feighery R, O J, Ravi N, Kelleher D, et al. 708 IL-1B and SERPINA-3 Are Novel Markers of Aggressive Barrett’s Oesophagus Phenotype Using RNA Deep Sequencing Analysis. Gastroenterology. 2014;146:S-122.

51. Maag JLV, Fisher OM, Levert-Mignon A, Kaczorowski DC, Thomas ML, Hussey DJ, et al. Novel aberrations uncovered in Barrett’s esophagus and esophageal adenocarcinoma using whole transcriptome sequencing. Molecular Cancer Research. 2017;15:1558–69.

52. Zhang Y, Weh KM, Howard CL, Riethoven JJ, Clarke JL, Lagisetty KH, et al. Characterizing isoform switching events in esophageal adenocarcinoma. Mol Ther Nucleic Acids. 2022;29:749–68.

53. Kimchi ET, Posner MC, Park JO, Darga TE, Kocherginsky M, Karrison T, et al. Progression of Barrett’s Metaplasia to Adenocarcinoma Is Associated with the Suppression of the Transcriptional Programs of Epidermal Differentiation.

54. Ostrowski J, Mikula M, Karczmarski J, Rubel T, Wyrwicz LS, Bragoszewski P, et al. Molecular defense mechanisms of Barrett’s metaplasia estimated by an integrative genomics. J Mol Med. 2007;85:733–43.

55. Stairs DB, Nakagawa H, Klein-Szanto A, Mitchell SD, Silberg DG, Tobias JW, et al. Cdx1 and c-Myc foster the initiation of transdifferentiation of the normal esophageal squamous epithelium toward Barrett’s esophagus. PLoS One. 2008;3.

56. Silvers AL, Lin L, Bass AJ, Chen G, Wang Z, Thomas DG, et al. Decreased selenium-binding protein 1 in esophageal adenocarcinoma results from posttranscriptional and epigenetic regulation and affects chemosensitivity. Clinical Cancer Research. 2010;16:2009–21.

57. Di Pietro M, Lao-Sirieix P, Boyle S, Cassidy A, Castillo D, Saadi A, et al. Evidence for a functional role of epigenetically regulated midcluster HOXB genes in the development of Barrett esophagus. Proc Natl Acad Sci U S A. 2012;109:9077–82.

58. Wang Q, Ma C, Kemmner W. Wdr66 is a novel marker for risk stratification and involved in epithelial-mesenchymal transition of esophageal squamous cell carcinoma. BMC Cancer. 2013;13.

59. Hyland PL, Hu N, Rotunno M, Su H, Wang C, Wang L, et al. Global changes in gene expression of Barrett’s esophagus compared to normal squamous esophagus and gastric cardia tissues. PLoS One. 2014;9.

60. Cummings LC, Thota PN, Willis JE, Chen Y, Cooper GS, Furey N, et al. A nonrandomized trial of vitamin D supplementation for Barrett’s esophagus. PLoS One. 2017;12.

61. Kastelein F, Biermann K, Steyerberg EW, Verheij J, Kalisvaart M, Looijenga LHJ, et al. Aberrant p53 protein expression is associated with an increased risk of neoplastic progression in patients with Barrett’s oesophagus. Gut. 2013;62:1676–83.

62. Li J, Li M han, Wang T tian, Liu X ning, Zhu X ting, Dai Y zhang, et al. SLC38A4 functions as a tumour suppressor in hepatocellular carcinoma through modulating Wnt/β-catenin/MYC/HMGCS2 axis. Br J Cancer. 2021;125:865–76.

63. Shulgin AA, Lebedev TD, Prassolov VS, Spirin P V. Plasmolipin and Its Role in Cell Processes. Mol Biol. 2021;55:773.

64. Jadhav K, Zhang Y. Activating transcription factor 3 in immune response and metabolic regulation. Liver Res. 2017;1:96–102.

65. Maag JLV, Fisher OM, Levert-Mignon A, Kaczorowski DC, Thomas ML, Hussey DJ, et al. Novel aberrations uncovered in Barrett’s esophagus and esophageal adenocarcinoma using whole transcriptome sequencing. Molecular Cancer Research. 2017;15:1558–69.

66. Lamouille S, Xu J, Derynck R. Molecular mechanisms of epithelial-mesenchymal transition. Nat Rev Mol Cell Biol. 2014;15:178–96.

67. Li K, Duan P, He H, Du R, Wang Q, Gong P, et al. Construction of the Interaction Network of Hub Genes in the Progression of Barrett’s Esophagus to Esophageal Adenocarcinoma. J Inflamm Res. 2023;16:1533–51.

68. Qi W, Li R, Li L, Li S, Zhang H, Tian H. Identification of key genes associated with esophageal adenocarcinoma based on bioinformatics analysis. Ann Transl Med. 2021;9:1711–1711.

69. Nancarrow DJ, Clouston AD, Smithers BM, Gotley DC, Drew PA, Watson DI, et al. Whole genome expression array profiling highlights differences in mucosal defense genes in barrett’s esophagus and esophageal adenocarcinoma. PLoS One. 2011;6.

70. Zhang Q, Agoston AT, Pham TH, Zhang W, Zhang X, Huo X, et al. Acidic Bile Salts Induce Epithelial to Mesenchymal Transition via VEGF Signaling in Non-Neoplastic Barrett’s Cells. Gastroenterology. 2019;156:130–144.e10.

71. Yao C, Li Y, Luo L, Xiong Q, Zhong X, Xie F, et al. Identification of miRNAs and genes for predicting Barrett’s esophagus progressing to esophageal adenocarcinoma using miRNA-mRNA integrated analysis. PLoS One. 2021;16.

72. Kalatskaya I. Overview of major molecular alterations during progression from Barrett’s esophagus to esophageal adenocarcinoma. Ann N Y Acad Sci. 2016;1381:74–91.

73. Darlavoix T, Seelentag W, Yan P, Bachmann A, Bosman FT. Altered expression of CD44 and DKK1 in the progression of Barrett’s esophagus to esophageal adenocarcinoma. Virchows Archiv. 2009;454:629–37.

74. Feith M, Stein HJ, Mueller J, Siewert JR. Malignant degeneration of Barrett’s esophagus: the role of the Ki-67 proliferation fraction, expression of E-cadherin and p53. 2004.

75. Falkenback D, Nilbert M, Öberg S, Johansson J. Prognostic value of cell adhesion in esophageal adenocarcinomas. Diseases of the Esophagus. 2008;21:97–102.

76. Paulson TG, Galipeau PC, Oman KM, Sanchez CA, Kuhner MK, Smith LP, et al. Somatic whole genome dynamics of precancer in Barrett’s esophagus reveals features associated with disease progression. Nat Commun. 2022;13.

77. Pinto R, Hauge T, Jeanmougin M, Pharo HD, Kresse SH, Honne H, et al. Targeted genetic and epigenetic profiling of esophageal adenocarcinomas and non-dysplastic Barrett’s esophagus. Clin Epigenetics. 2022;14.

78. Redston M, Noffsinger A, Kim A, Akarca FG, Rara M, Stapleton D, et al. Abnormal TP53 Predicts Risk of Progression in Patients With Barrett’s Esophagus Regardless of a Diagnosis of Dysplasia. Gastroenterology. 2022;162:468–81.

79. Januszewicz W, Pilonis ND, Sawas T, Phillips R, O’Donovan M, Miremadi A, et al. The utility of P53 immunohistochemistry in the diagnosis of Barrett’s oesophagus with indefinite for dysplasia. Histopathology. 2022;80:1081–90.

80. Kalatskaya I. Overview of major molecular alterations during progression from Barrett’s esophagus to esophageal adenocarcinoma. Ann N Y Acad Sci. 2016;1381:74–91.

81. Kastelein F, Biermann K, Steyerberg EW, Verheij J, Kalisvaart M, Looijenga LHJ, et al. Aberrant p53 protein expression is associated with an increased risk of neoplastic progression in patients with Barrett’s oesophagus. Gut. 2013;62:1676–83.

82. Li S, Hoefnagel SJM, Krishnadath KK. Molecular Biology and Clinical Management of Esophageal Adenocarcinoma. Cancers 2023, Vol 15, Page 5410. 2023;15:5410.

83. Redston M, Noffsinger A, Kim A, Akarca FG, Rara M, Stapleton D, et al. Abnormal TP53 Predicts Risk of Progression in Patients With Barrett’s Esophagus Regardless of a Diagnosis of Dysplasia. Gastroenterology. 2022;162:468.

84. Eluri S, Brugge WR, Daglilar ES, Jackson SA, Styn MA, Callenberg KM, et al. The presence of genetic mutations at key loci predicts progression to esophageal adenocarcinoma in Barrett’s esophagus. American Journal of Gastroenterology. 2015;110:828–34.

85. Iyer PG, Codipilly DC, Chandar AK, Agarwal S, Wang KK, Leggett CL, et al. Prediction of Progression in Barrett’s Esophagus Using a Tissue Systems Pathology Test: A Pooled Analysis of International Multicenter Studies. Clinical Gastroenterology and Hepatology. 2022. 10.1016/j.cgh.2022.02.033.

86. Codipilly DC, Krishna Chandar A, Wang KK, Katzka DA, Goldblum JR, Thota PN, et al. Wide-area transepithelial sampling for dysplasia detection in Barrett’s esophagus: a systematic review and meta-analysis. Gastrointest Endosc. 2022;95:51–59.e7.

87. Ross-Innes CS, Chettouh H, Achilleos A, Galeano-Dalmau N, Debiram-Beecham I, MacRae S, et al. Risk stratification of Barrett’s oesophagus using a non-endoscopic sampling method coupled with a biomarker panel: a cohort study. Lancet Gastroenterol Hepatol. 2017;2:23–31.

88. Panda A, Bhanot G, Ganesan S, Bajpai M. Gene expression in barrett’s esophagus cell lines resemble esophageal squamous cell carcinoma instead of esophageal adenocarcinoma. Cancers (Basel). 2021;13:5971.

89. Odze RD. Update on the Diagnosis and Treatment of Barrett Esophagus and Related Neoplastic Precursor Lesions. Arch Pathol Lab Med. 2007;132:1577–85.

90. Kaul V, Gross S, Corbett FS, Malik Z, Smith MS, Tofani C, et al. Clinical utility of wide-area transepithelial sampling with three-dimensional computer-assisted analysis (WATS3D) in identifying Barrett’s esophagus and associated neoplasia. Diseases of the Esophagus. 2020;33.

